# Phenotype-based targeted treatment of SGLT2 inhibitors and GLP-1 receptor agonists in type 2 diabetes

**DOI:** 10.1101/2023.08.04.23293636

**Authors:** Pedro Cardoso, Katie G. Young, Anand T.N. Nair, Rhian Hopkins, Andrew P McGovern, Eram Haider, Piyumanga Karunaratne, Louise Donnelly, Bilal A. Mateen, Naveed Sattar, Rury R. Holman, Jack Bowden, Andrew T. Hattersley, Ewan R. Pearson, Angus G. Jones, Beverley M. Shields, Trevelyan J. McKinley, John M. Dennis, The MASTERMIND consortium

**Affiliations:** University of Exeter Medical School. Address: Institute of Biomedical & Clinical Science, RILD Building, Royal Devon & Exeter Hospital, Barrack Road, Exeter EX2 5DW, UK; University of Dundee. Address: Division of Molecular & Clinical Medicine, Ninewells Hospital and Medical School, University of Dundee, Dundee DD1 9SY, UK; University College London, Institute of Health Informatics. Address: University College London, 222 Euston Rd, London NW1 2DA, London, UK; Institute of Cardiovascular and Medical Sciences, University of Glasgow, Glasgow G12 8TA, UK; Diabetes Trials Unit, Oxford Centre for Diabetes, Endocrinology and Metabolism, University of Oxford. Oxford NIHR Biomedical Research Centre, Churchill Hospital, Oxford. Address: Diabetes Trials Unit, Oxford Centre for Diabetes, Endocrinology and Metabolism, University of Oxford, Oxford, UK

**Keywords:** Bayesian non-parametric modelling, heterogeneous treatment effects, precision medicine, type 2 diabetes, SGLT2-inhibitors, GLP1-receptor agonists

## Abstract

A precision medicine approach in type 2 diabetes (T2D) could enhance targeting specific glucose-lowering therapies to individual patients most likely to benefit. We utilised Bayesian non-parametric modelling to develop and validate an individualised treatment selection algorithm for two major T2D drug classes, SGLT2-inhibitors (SGLT2i) and GLP1-receptor agonists (GLP1-RA). The algorithm is designed to predict differences in 12-month glycaemic outcome (HbA_1c_) between the 2 therapies, based on routine clinical features of 46,394 people with T2D in England (27,319 for model development, 19,075 for hold-out validation), with additional external validation in 2,252 people with T2D from Scotland. Routine clinical features, including sex (with females markedly more responsive to GLP1-RA), were associated with differences in glycaemic outcomes. Our algorithm identifies clearly delineable subgroups with reproducible ≥5mmol/mol HbA_1c_ benefits associated with each drug class. Moreover, we demonstrate that targeting the therapies based on predicted glycaemic response is associated with improvements in short-term tolerability and long-term risk of new-onset microvascular complications. These results show that precision medicine approaches to T2D can facilitate effective individualised treatment selection, and that use of routinely collected clinical features could support low-cost deployment in many countries.

## Introduction

A precision medicine approach in type 2 diabetes (T2D) would aim to target specific glucose-lowering therapies to individual patients most likely to benefit—for a detailed review of the state of the field, see Dennis, 2020 [1]. Current stratification in T2D treatment guidelines involves preferential prescribing of two major drug classes, SGLT2i-inhibitors (SGLT2i) and GLP1-receptor agonists (GLP1-RA), to subgroups of people with or at high-risk of cardiorenal disease [2]. Evidence informing these recommendations comes from average treatment effect estimates derived from placebo-controlled cardiovascular and renal outcome trials, which have predominantly recruited participants with advanced atherosclerotic cardiovascular risk or established cardiovascular disease [3, 4]. Consequently, there is limited evidence on the benefits of SGLT2i and GLP1-RA for individuals in the broader T2D population and, given the lack of head-to-head trials, of the relative efficacy of the two drug classes for individual patients.

Recent studies have demonstrated a clear potential for a precision medicine approach based on glycaemic response, with the TRIMASTER crossover trial establishing a greater efficacy of SGLT2i compared to another major drug class, DPP4-inhibitors (DPP4i), in those with better renal function, and a greater efficacy of thiazolidinedione therapy compared with DPP4i in those with obesity (BMI >L30Lkg/m) compared to those without obesity [5]. Given these findings, a validated prediction model to support individualised treatment selection has recently been developed for SGLT2i compared with DPP4i therapy using observational and clinical trial data [6]. For GLP1-RA, although recent studies have identified robust heterogeneity in treatment response based on pharmacogenetic markers and markers of insulin secretion [7, 8], the influence of these markers on relative differences in clinical outcomes compared with other drug classes, and therefore their utility for targeting treatment, has not previously been assessed.

Given the lack of evidence to support targeted treatment of SGLT2i compared with GLP1-RA therapies, we aimed to develop and validate a prediction model to provide individualised estimates of differences in 12-month glycaemic (HbA_1c_) outcomes for the two drug classes. We developed a model using the recently proposed Bayesian Causal Forest (BCF) structure, designed to explicitly identify and predict conditional average treatment effects (CATE), representing differential effects of the two drug classes on HbA1c outcome conditional on the clinical characteristics of individual patients [9]. The BCF framework also minimises confounding from indication bias and allows for flexibility in defining model structure and outputs. Our approach is based on readily-available and routinely collected clinical features, supporting potential low-cost deployment in most countries. We also evaluated the downstream impacts of targeting therapy based on the glycaemic response on secondary outcomes of weight change, tolerability, and longer-term risk of new-onset microvascular complications, macrovascular complications, and adverse kidney events.

## Results

We identified 112,274 study-eligible non-insulin treated people with T2D initiating SGLT2i (n=84,193) or GLP1-RA (n=28,081) for the first time between January 2013 and October 2020 in UK general practice, accessed from Clinical Practice Research Datalink (CPRD) (sFlowchart 1). The mean age of participants was 58.2 (SD=10.9) years, 66,248 (59%) were men, and 88,174 (79%) were of white ethnicity. Baseline clinical characteristics by initiated drug class are listed in Table 1. For the development of the 12-month HbA_1c_ response treatment selection model, individuals with a measured HbA_1c_ outcome were randomly split 60:40 into development (n=31,346) and validation (n=20,865) cohorts (sFlowchart 1; Baseline characteristics by cohort: sTable 1A-B). Mean unadjusted 12-month HbA1c response (change from baseline in HbA1c) was -12.0 (SD 15.3) mmol/mol for SGLT2i and -11.7 (SD 17.6) mmol/mol for GLP1-RA. Specific cohorts were defined for secondary outcomes to maximise the number of included patients for each analysis (sFlowchart 2).

**Table 1.**
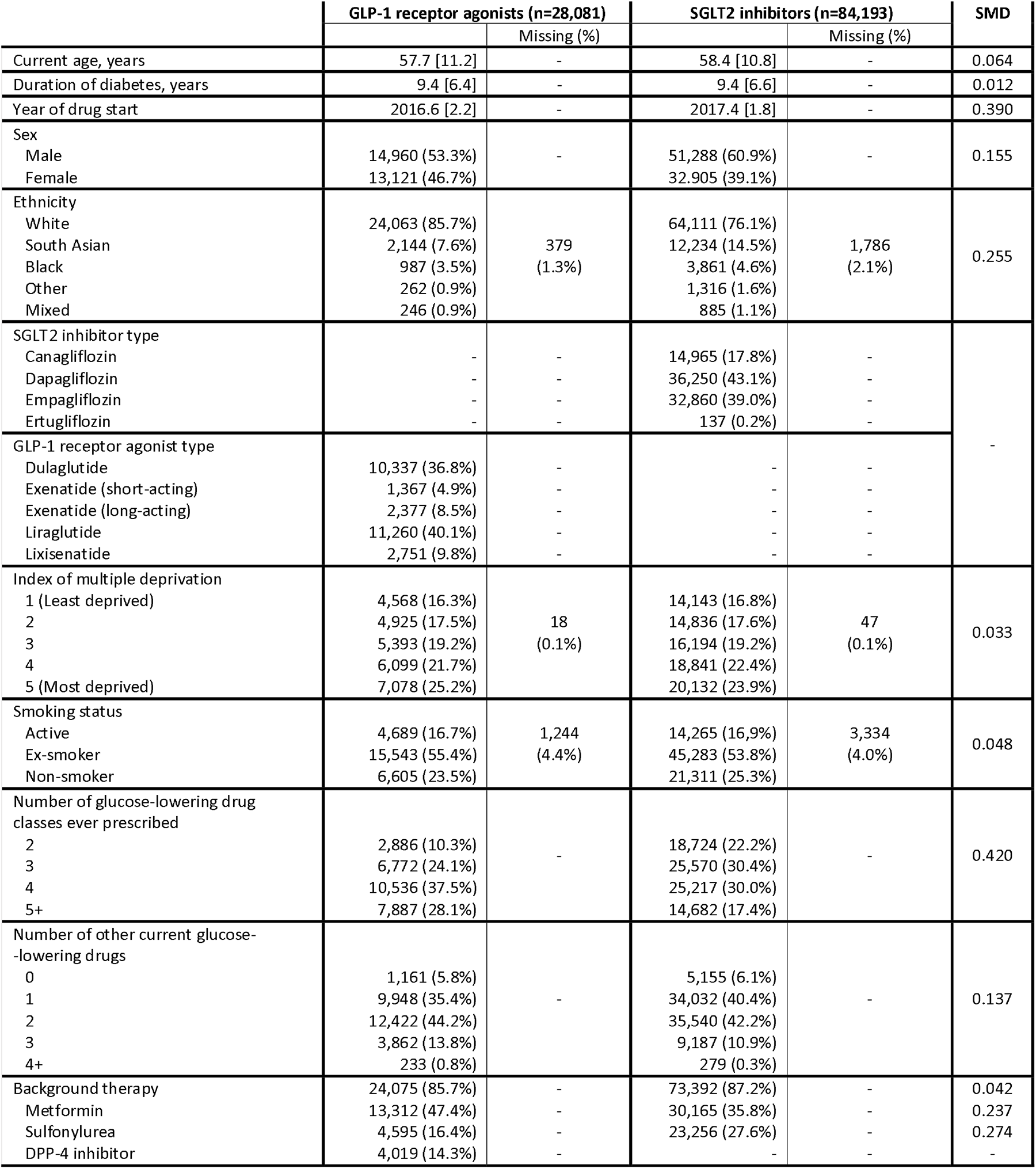
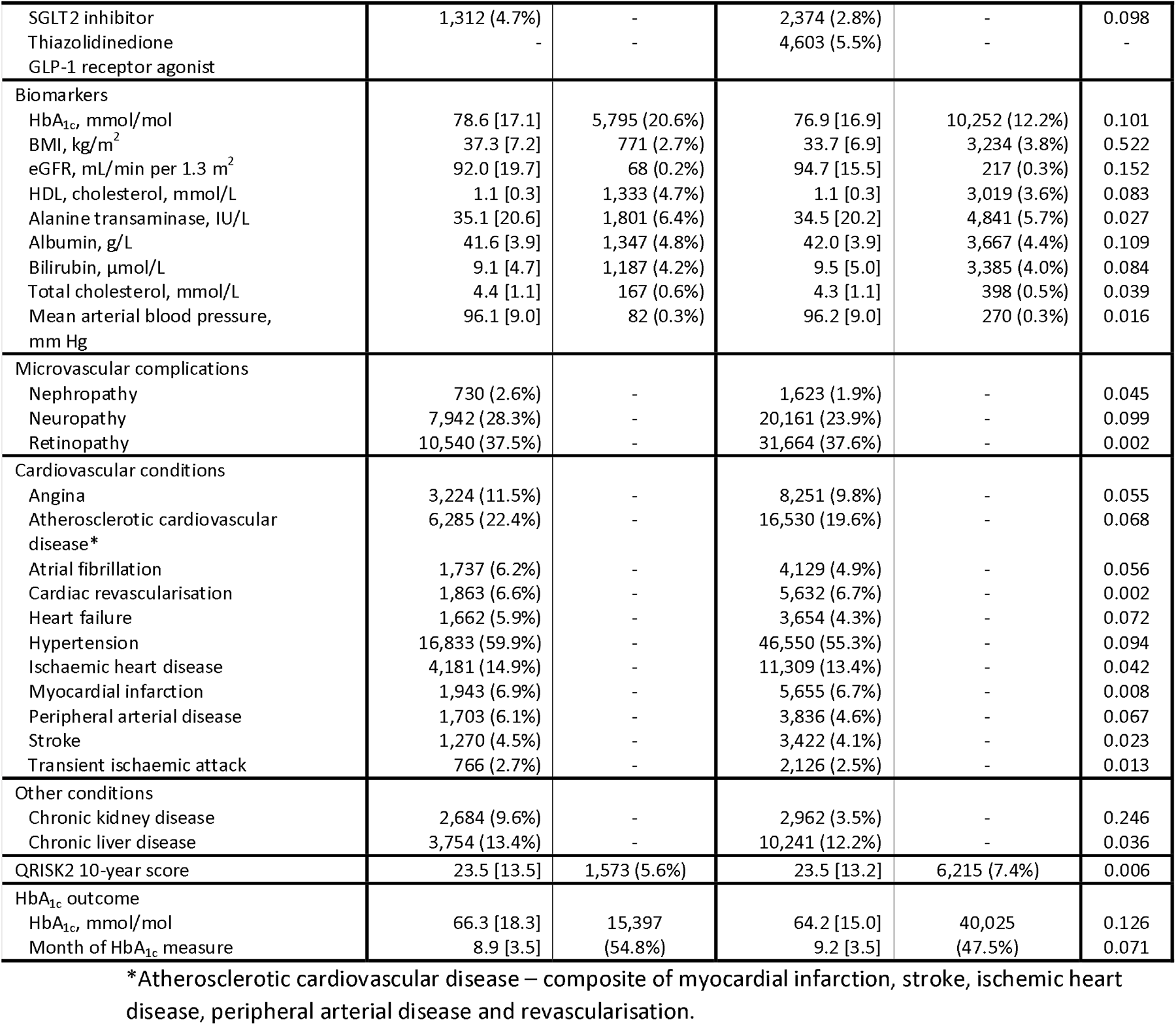
Baseline clinical characteristics of patients initiating GLP-1 receptor agonists and SGLT2-inhibitors from the UK Clinical Practice Research Datalink. Data are mean [SD] and number (%). SMD: Standardised mean difference (≥0.1 is a common metric for a meaningful imbalance between treatment groups).

### Model development

After variable selection [9] (sFig. 1), the BCF model identified multiple clinical factors predictive of HbA_1c_ response with SGLT2i (the reference drug class in the model) henceforth referred to as the prognostic factors, and multiple factors predictive of differential HbA_1c_ response with GLP1-RA compared to SGLT2i therapy (henceforth referred to as the differential factors —Table 2). The final BCF model was fitted to 27,319 (87.2%) individuals with complete data for all selected clinical factors. Overall model fit and performance statistics for predicting achieved HbA_1c_ outcome in internal validation for both the development and hold-out cohorts are reported in sTable 2. Model predictions were similar whether or not we incorporated a propensity score into the BCF model (as recommended by Hahn et al. [10]) (sFig. 2). Therefore, we removed the propensity score covariate from the final model to support easier future implementation of the model within clinical practice.

**Table 2.** Baseline clinical features included in the treatment selection algorithm after variable selection.

| **Features predictive of HbA1c outcome with SGLT2i** | **Features predictive of differential HbA1c outcome with GLP1-RA compared to SGLT2i** |
| --- | --- |
| Current age, years | Current age, years |
| Duration of diabetes, years | Sex |
| Number of glucose-lowering drug classes ever prescribed | Number of other current glucose-lowering drugs |
| Number of other current glucose-lowering drugs | HbA<sub>1c</sub>, mmol/mol |
| HbA<sub>1c</sub>, mmol/mol | BMI, kg/m<sup>2</sup> |
| eGFR, mL/min per 1.3 m<sup>2</sup> | eGFR, mL/min per 1.3 m<sup>2</sup> |
| Alanine transaminase, IU/L | Heart Failure |
| Peripheral arterial disease | Ischaemic heart disease |
|  | Neuropathy |
|  | Peripheral arterial disease |
|  | Retinopathy |

In the development cohort, the predicted CATE was a 0.1 (95%CI -0.3;0.5) mmol/mol benefit with GLP1-RA over SGLT2i, suggesting similar average efficacy of both therapies. However, across individuals, there was marked heterogeneity in predicted CATE (Fig. 1A), with the model predicting an average HbA_1c_ benefit on SGLT2i therapy for 13,110 (48%) individuals and on GLP1-RA for 14,209 (52%) individuals. 4,787 (17.5%) of the development cohort had a predicted HbA1c benefit >3 mmol/mol (3mmol/mol is used widely as minimally important difference in clinical trials) with SGLT2i over GLP1-RA, and 5,551 (20.3%) had a predicted HbA_1c_ benefit >3 mmol/mol with GLP1-RA over SGLT2i.

**Fig. 1:**
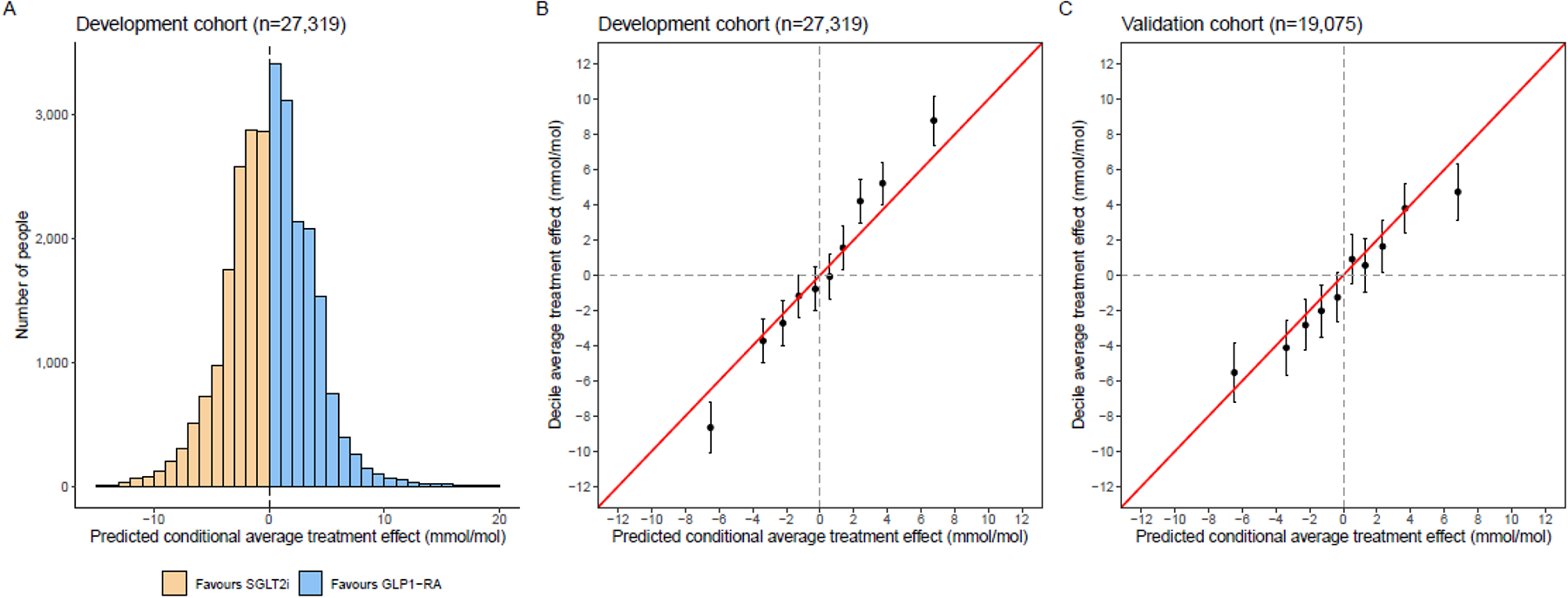
Predicted conditional average treatment effects and model calibration. (A) Distribution of conditional average treatment effect (CATE) estimates for SGLT2-inhibitors vs. GLP-1 receptor agonists in the CPRD development cohort; negative values reflect a predicted HbA1c treatment benefit on SGLT2-inhibitors and positive values reflect a predicted treatment benefit on GLP-1 receptor agonists. (B) Calibration between average treatment effects (ATE) and predicted CATE estimates, by decile of predicted CATE in the development cohort. (C) Calibration of CATE estimates in the validation cohort. ATE estimates are adjusted for all the variables used in the treatment selection model (see Methods).

### Model calibration

Calibration by decile of model-predicted CATE estimates was good in the development cohort (n=27,319; Fig. 1B), the hold-back CPRD validation cohort (n=19,075, Fig. 1C), and in propensity-matched cohorts (sFig. 3). In practice, true CATE estimates are unobserved since a single individual receives only one therapy, meaning the counterfactual outcome they would have had on the alternative therapy is unobserved. Therefore, for each decile calibration was based on comparing mean predicted CATE estimates to the mean differences in outcome HbA1c of individuals receiving SGLT2i compared to GLP1-RA—for full details, see Methods.

We also evaluated model performance in an external Scottish cohort—Tayside & Fife (n=2,252 [1,837 initiating SGLT2i, 415 initiating GLP1-RA]; Baseline characteristics: sTable 1C). A similar distribution of predicted CATE to CPRD was observed (Extended Fig. 1A). Due to the smaller cohort size, calibration was assessed by quintile of predicted CATE, with a clear difference between upper (favouring GLP1-RA) and lower (favouring SGLT2i) quintiles, but modest calibration in middle quintiles (Extended Fig. 1B). Among 81 (3.6%) individuals with a model-predicted HbA_1c_ benefit >5 mmol/mol for SGLT2i over GLP1-RA, there was a 7.4 mmol/mol (95%CI 0.1;14.8) benefit for SGLT2i (Extended Fig. 1C). In contrast, among 150 (6.7%) individuals with a model-predicted HbA_1c_ benefit >5 mmol/mol for GLP1-RA over SGLT2i, there was a 5.6 mmol/mol (95%CI -0.9;12.1) benefit on GLP1-RA.

### Model interpretability

Stratifying the combined development and validation cohorts with complete predictor data (n=46,394) into subgroups defined by predicted CATE, there were clear differences in clinical characteristics, with those having a greater predicted HbA1c benefit with GLP1-RA over SGLT2i being predominantly female and older, with lower baseline HbA_1c_, eGFR and BMI (Fig. 2, sTable 1E). SGLT2i were predicted to have a greater HbA_1c_ benefit over GLP1-RA for 32% of those with baseline HbA_1c_ levels <64, 39% 64-75, 54% 75-86, and 67% ≥86 mmol/mol. Given their average older age, those with the greatest (>5 mmol/mol) predicted benefit on GLP1-RA also had a higher prevalence of microvascular complications (75.2% versus 40.8%), cardiovascular disease (36.5% versus 15.8%), chronic kidney disease (25.1% versus 0.9%), and heart failure (8.8% versus 4.2%) (sTable 1E). An evaluation of relative variable importance identified the number of other current glucose-lowering drugs (a higher number of concurrent therapies favouring SGLT2i as the optimal treatment), sex, current age, and to a lesser extent BMI and HbA_1c_ as the most influential predictors (relative importance ≥3%). In contrast, microvascular complications and cardiovascular comorbidities had very modest effects on differential response (Extended Fig. 2).

**Fig. 2:**
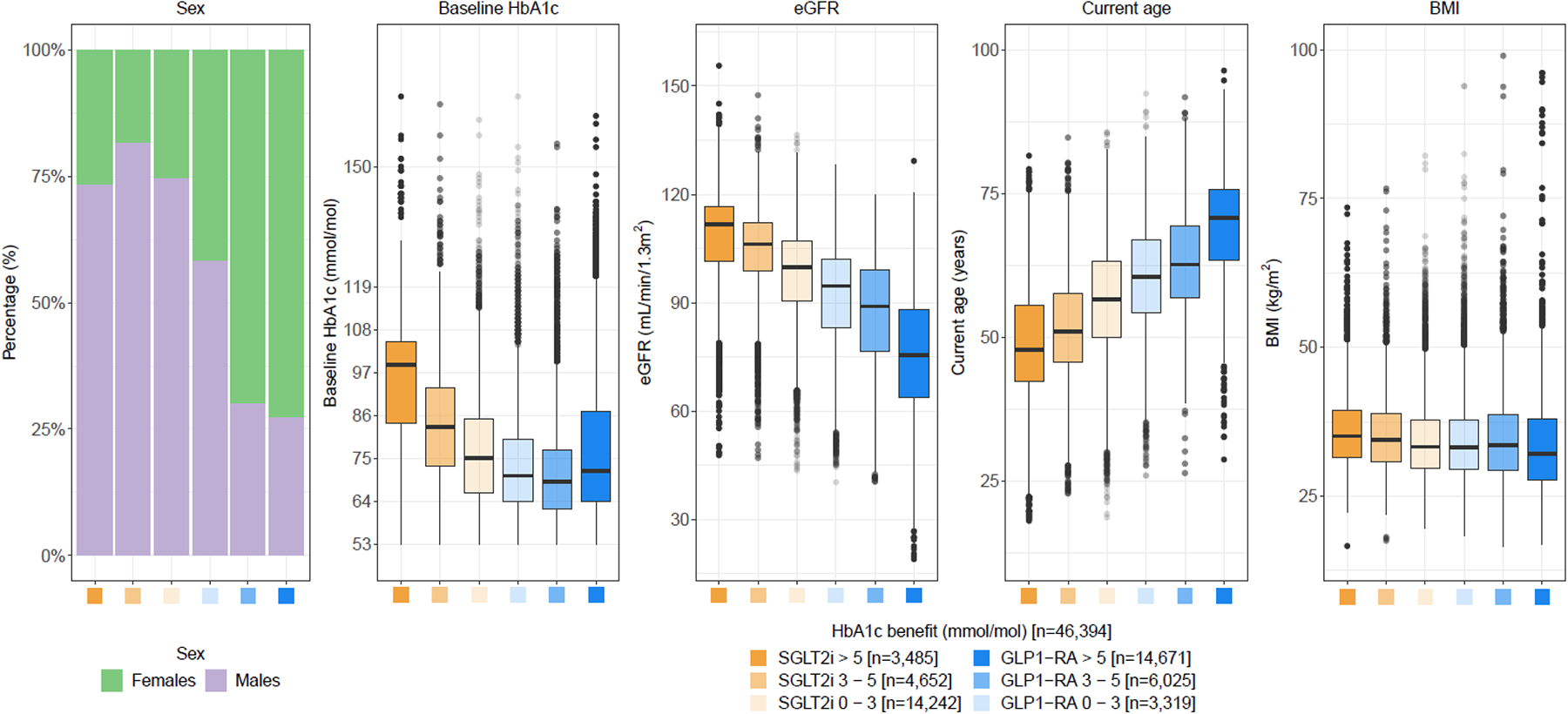
Distributions of major clinical characteristics predicting differential HbA1c outcome with SGLT2i and GLP1-RA. Distributions of key differential clinical characteristics in the combined development and validation cohorts (n=46,394 with complete predictor data) for subgroups defined by predicted HbA_1c_ outcome differences: SGLT2i benefit ≥5 mmol/mol, 3–5 mmol/mol and 0–3 mmol/mol, GLP1-RA benefit ≥5 mmol/mol, 3–5 mmol/mol and 0–3 mmol/mol.

### Replication of sex differences in glycaemic response in clinical trials

Whilst previous analyses of clinical trials and observational data for SGLT2i have shown a greater HbA_1c_ response in males compared to females, which we additionally reproduced in Tayside & Fife (Extended Fig. 3), sex differences in GLP1-RA response have not been clearly established. Here, we focused on individual-level randomised clinical trial data of GLP1-RA from the HARMONY programme (Liraglutide [n=389] and Albiglutide [n=1,682]), [11], the PRIBA prospective cohort study (Liraglutide [n=397], exenatide [n=223]) [7], and Tayside & Fife (n=415). Baseline characteristics for the cohorts are reported in sTable 1C-D. Across all studies, there was consistent evidence of a greater baseline HbA_1c_ adjusted glycaemic response in females versus males; this was most marked for Liraglutide in the HARMONY 7 trial [11] where a 4.4 (95%CI 2.2;6.3) mmol/mol greater response in females than males was observed.

### Effect of targeting therapy based on predicted HbA_1c_ outcome on other short- and long-term outcomes

To assess the potential associations of targeting treatment based on predicted HbA_1c_ benefit with other short- and long-term outcomes, we divided the study population into six subgroups based on clinically relevant differences in predicted CATE (HbA1c differences of 0-3, 3-5 and >5 mmol/mol between therapies). We then compared average differences in each short and long-term outcome in individuals receiving SGLT2i compared to GLP1-RA within each subgroup, for adjusted (Fig. 3), propensity-matched (sFig. 4), and double robust adjusted models (sFig. 5). Specific subpopulations were defined for each short-term outcome based on the availability of observed outcome data (12-month HbA_1c_ change from baseline [to evaluate absolute response] n=87,835; 12-month weight change n=41,728; treatment discontinuation within 6 months [a proxy for tolerability] n=77,741) (sFlowchart 2). Longer-term outcomes were evaluated up to 5 years from drug initiation, excluding individuals with a history of cardiovascular disease or chronic kidney disease for major adverse cardiovascular event (MACE), heart failure, and adverse kidney (composite of ≥40% decline in eGFR or kidney failure [12]) outcomes (n=52,052) and individuals with a history of retinopathy, neuropathy and nephropathy for microvascular outcome (n=34,524). (sFlowchart 2).

**Fig. 3:**
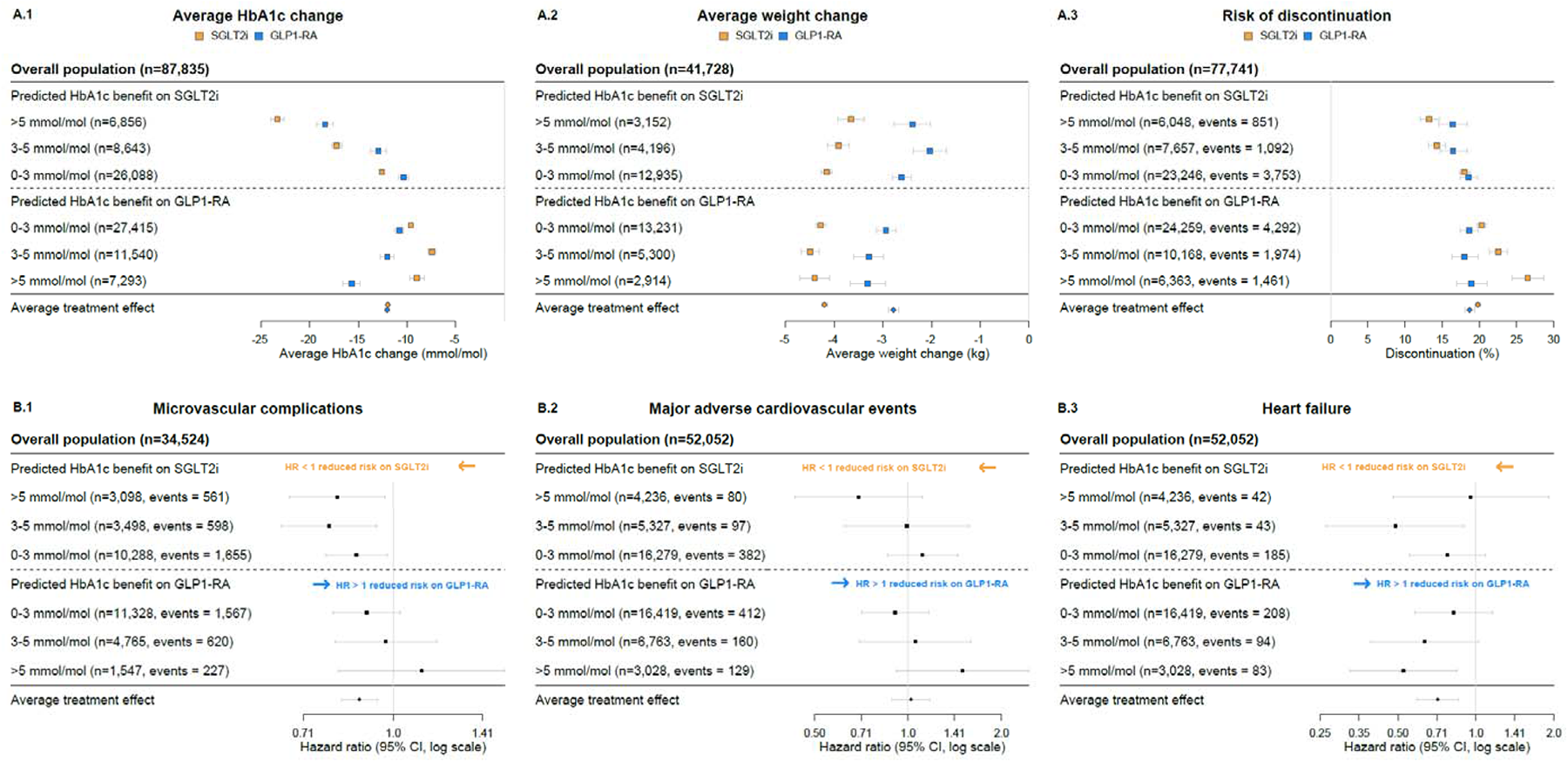
Differences in short-term and long-term clinical outcomes with SGLT2i and GLP1-RA for subgroups defined by predicted HbA1c response differences. (A.1) 12-month HbA1c change from baseline. (A.2) 12-month weight change. (A.3) 6-month risk of discontinuation. (B.1) Hazard ratios for 5-year risk of new-onset microvascular complications (retinopathy, nephropathy or neuropathy). (B.2) Hazard ratios for 5-year relative risk of major adverse cardiovascular events (MACE). (B.3) Hazard ratios for 5-year risk of heart failure. Hazard ratios represent the relative risk for those treated with GLP1-RA in comparison to SGLT2i therapy, with a value under 1 favouring SGLT2i therapy. Data underlying the Figure are reported in sTable 3.

For HbA_1c_ change from baseline, of the 6,856 individuals (7.8%) with a predicted HbA_1c_ benefit on SGLT2i of ≥5 mmol/mol, those who received SGLT2i had on average a 23.3 mmol/mol (95%CI 22.6;24.0) reduction in HbA_1c_ and those who received GLP1-RA had on average an 18.4 mmol/mol (95%CI 17.6;19.3) reduction in HbA_1c_ (Fig. 3A.1). In contrast, 7,293 individuals (8.3%) with a predicted HbA_1c_ benefit on GLP1-RA of ≥5 mmol/mol, those receiving GLP1-RA had on average a 15.7 mmol/mol (95%CI 14.8;16.6) reduction in HbA_1c_, and those receiving SGLT2i had on average a 9.0 mmol/mol (95%CI 8.2;9.7) reduction in HbA_1c_. Similar differences between subgroups were seen when stratified by major UK self-reported ethnicity groups (White and Non-White [South Asian, Black, Other or Mixed]) (Extended Fig. 4), and by history of cardiovascular disease (sFig. 10).

Observed weight change was consistently greater for individuals treated with SGLT2i compared to GLP1-RA across all subgroups (Fig. 3A.2). Short-term discontinuation was lower in those treated with the drugs predicted to have the greatest glycaemic benefit by the model, mainly reflecting differences in SGLT2 discontinuation across predicted levels of differential glycaemic response (Fig. 3A.3). Relative risk of new-onset microvascular complications also varied by subgroup, with a lower risk with SGLT2i versus GLP1-RA only in subgroups predicted to have a glycaemic benefit with SGLT2i (Fig. 3B.1). HRs for the risk of new-onset MACE were similar overall (HR 1.02 [95%CI 0.89;1.18]) and by subgroup (Fig. 3B.2). HRs for the risks of both new-onset heart failure and adverse kidney outcomes were lower with SGLT2i (heart failure HR 0.71 [95%CI 0.59;0.85]; CKD HR 0.41 [95%CI 0.30;0.56]) with no clear evidence of a difference by subgroup (Fig. 3B.3, sFig. 6C).

### Comparison of model predictions with our previously published treatment selection model for SGLT2i and DPP4i therapies

In n=82,933 eligible patients, predictions for HbA_1c_ response with SGLT2i from the SGLT2i v GLP1-RA treatment selection model were highly concordant (R > 0.92) with those from our recently published SGLT2i versus DPP4i treatment selection model, which was developed in an independent UK primary care cohort [6] (sFig. 7). Estimating differential HbA_1c_ responses using both models in our study population with complete data suggested SGLT2i is the predicted optimal therapy for HbA_1c_ in 48.2% (n=39,975) of individuals, GLP1-RA the predicted optimal therapy in 51.3% (n=42,519), and DPP4i the optimal therapy for only 0.5% (n=439).

### Prototype treatment selection model

A prototype treatment selection model app (provided for research purposes only) providing individualised predictions of differences (with uncertainty) in HbA1c outcomes is available at: https://pml204.shinyapps.io/SGLT2_GLP1_calculator/

## Discussion

We have developed a novel treatment selection algorithm using state-of-the-art Bayesian methods to predict differences in one-year glycaemic outcomes for SGLT2i and GLP1-RA and validated the algorithm in a hold-out cohort and an independent external validation cohort. Our individualised precision medicine approach shows glycaemic response-based targeting of these two major drug classes in people with T2D can not only optimise glycaemic control, but is also associated with improved tolerability and reduced risk of new-onset microvascular complications. In contrast, we found limited evidence for heterogeneity in other clinical outcomes, with overall equipoise between the two therapies for new-onset MACE and a clear overall benefit with SGLT2i over GLP1-RA for new-onset heart failure and adverse kidney outcomes (≥40% eGFR decline or renal failure). Predictions are based on individual patient-level routine clinical characteristics, meaning the model can be deployed in most countries worldwide without the need for additional testing.

Our approach differs from notable recent studies that have attempted to sub-classify people with T2D or used dimensionality reduction to represent T2D heterogeneity [13, 14, 15]. Whilst these alternative approaches can provide important insight into underlying heterogeneities of T2D patient characteristics, they, by definition, lose information about the specific characteristics of individual patients, meaning they are sub-optimal for accurately predicting the treatment or disease progression outcomes for individuals [16]. If subclassification approaches based on clinical features are to have potential clinical utility, they will need to be updated over time as an individual’s phenotype evolves [17]. In contrast, our ‘outcomes-based’ approach enables the prediction of optimal therapy when a treatment decision is made, uses the specific information available for a patient at that point in time and avoids sub-classification.

Although BCF models are only causal under specific assumptions [18], they can provide insights into differences in the underlying mechanisms of action of GLP1-RA and SGLT2i, and the clinical utility of these differences. The strongest predictor of a differential glycaemic response was the number of currently prescribed glucose-lowering therapies, which is a likely proxy of the degree of diabetes progression (and, therefore, underlying beta cell failure) of an individual. A plausible biological explanation for the relevance of this proxy is that an attenuated GLP1-RA response has been observed in individuals with more severe diabetes; previous clinical studies have identified that markers of beta cell failure (including longer diabetes duration, insulin co-treatment and lower fasting c-peptide) are associated with a lower glycaemic response to GLP1-RA [7], with no evidence of differences for SGLT2i [19]. The favouring of GLP1-RA over SGLT2i in females is novel but is supported by our trial validation and recent pharmacokinetic data demonstrating higher circulating GLP1-RA drug concentrations and, consequently, greater HbA1c reduction in females compared with males [20]. For SGLT2i, increased urinary glucose excretion likely explains the greater relative glycaemic efficacy with higher baseline HbA_1c_ and eGFR, which in concordance with our analysis, has been previously demonstrated in trial data [6]. Of note, the comorbidities included in the final model had modest effects on HbA1c and are likely to be proxy measures of factors underlying differential response to these therapies.

Our study represents the second application of our novel validation framework for precision medicine models, which, in the absence of true observed outcomes (for an individual patient on one therapy, the counterfactual outcome they would have had on an alternative therapy cannot be observed [21]), evaluates accuracy in subgroups defined by predicted CATE. The previous study developed a treatment selection model for SGLTi2 versus DPP4i therapy in a separate dataset, with models developed in primary care data validating well in head-to-head clinical trial datasets. Although this demonstrated marked heterogeneity in the relative glycaemic outcome, most (84%) individuals had a greater glycaemic reduction with SGLT2i. In contrast, this GLP1-RA/SGLT2i model shows even greater heterogeneity in treatment effects but with equipoise on average treatment effects between the two therapies (52% favouring GLP1-RA). Furthermore, we demonstrate that optimising therapy based on predicted glycaemic response may lower microvascular complication risk, a finding concordant with evidence from the UKPDS study on the importance of good glycaemic control to lower the risk of microvascular disease [22, 23].

Further developments to this model could include the incorporation of non-routine and pharmacogenetic markers (recently identified for GLP1-RA) [8], and additional glucose-lowering drug classes, in particular, off-patent sulfonylureas and pioglitazone to support the deployment of the algorithm in lower-income countries where the availability of newer medications may be limited. Assessment of semaglutide, a GLP-1RA with potent glycaemic effect excluded here due to low numbers prescribed during the period of data availability, and tirzepatide, a dual GIP and GLP-1 receptor agonist not currently available in the UK, is an important area for future research as our model may benefit from recalibration for these newer therapies. Although our ethnicity-specific validation suggests good performance in people of non-white ethnicity, setting and ethnicity-specific validation and optimisation would also improve future clinical utility. Given the possibility of selection bias due to non-random treatment assignment, validation in a dataset where individuals were randomised to therapy would further strengthen the evidence for model deployment. However, few active comparator trials of these two therapies have been conducted [24], and to our knowledge, none are available for data-sharing. Ultimately, research, likely in even larger datasets, is needed on whether individualised models for other short-and-long term outcomes beyond glycaemia, particularly cardiorenal disease, can further improve current prescribing approaches [25].

Finally, a limitation of our study is that despite being state-of-the-art and with a key advantage of allowing estimation of predictions with uncertainty, and so facilitating more transparent evaluation, the Bayesian causal forest methods we applied are subject to ongoing development in several key areas such as variable selection [9, 26], scalability, and handling of missing data [27].

In conclusion, our study demonstrates a clear potential for targeted prescribing of GLP1-RA and SGLT2i to individual people with T2D based on their clinical characteristics to improve glycaemic outcomes, tolerability, and risk of microvascular complications. This provides an important advance on current T2D guidelines, which only recommend preferentially prescribing these therapies to individuals with, or at high risk of, cardiorenal disease, with no clear evidence to choose between the two drug classes. Precision T2D prescribing based on routinely-available characteristics has the potential to lead to more informed and evidence-based decisions on treatment for people with T2D worldwide in the near future.

## Methods

### Study population

Patients with type 2 diabetes initiating SGLT2i and GLP1-RA therapies between 1^st^ January 2013 and 31^st^ October 2020 were identified in the UK Clinical Practice Research Datalink (CPRD) Aurum dataset [28], following our previously published cohort profile [29] (see GitHub: Exeter-Diabetes/CPRD-Codelists for the codelists used). We excluded patients that were prescribed either therapy as first-line treatment (not recommended in UK guidelines) [30], those co-treated with insulin, and those with a diagnosis of end-stage renal disease (ERSD) (sFlowchart 1). Due to the low numbers of eligible patients we also excluded individuals initiating the GLP1-RA semaglutide (n=784 study eligible with outcome HbA1c recorded)) [31]. This final CPRD cohort was randomly split 60:40 into development and hold-back validation sets, maintaining the proportion of individuals receiving SGLT2i and GLP1-RA in each set.

### Additional cohorts

The same eligibility criteria were applied to define an independent population in Scotland for model validation (SCI-Diabetes [Tayside & Fife] [32], containing longitudinal observational data on biochemical investigations and prescriptions). To assess reproducibility of differences in HbA_1c_ response by sex with GLP1-RA therapy, we accessed individual-level data on participants initiating the GLP1-RAs Albiglutide and Liraglutide in the HARMONY clinical trial programme, an international randomised placebo-controlled trial designed to evaluate the cardiovascular benefit of Albiglutide with type 2 diabetes [11], and the PRIBA prospective cohort study (United Kingdom 2011-2013) [7], designed to test whether individuals with low insulin secretion have lesser glycaemic response to incretin-based treatments.

### Outcomes

The primary outcome was achieved HbA_1c_ at twelve months post-drug initiation for individuals who remained on unchanged glucose-lowering therapy. Given the variability in the timing of follow-up testing in UK primary care, this outcome was defined as the closest eligible HbA_1c_ value to 12 months (within 3–15 months) after initiation to maximise sample size. To allow for potential differential effects of follow-up duration on HbA_1c_, we included an additional covariate to capture the month post-initiation that the HbA_1c_ value was recorded.

Secondary outcomes comprised short-term outcomes: change in weight 12 months after initiation (closest recorded weight to 12 months, within 3–15 months), and, as a proxy for drug tolerability, treatment discontinuation within 6 months of drug initiation (as such short-term discontinuation is unlikely to be related to a lack of glycaemic response); and longer-term outcomes up to 5-years after initiation: new-onset major adverse cardiovascular events (MACE: composite of myocardial infarction, stroke and cardiovascular death); new-onset heart failure; and new-onset adverse kidney outcome (a drop of ≥40% in eGFR from baseline or reaching CKD stage 5 [12]); and new-onset microvascular complications (see sFlowchart 2).

### Predictors

Candidate predictors comprised — current age, duration of diabetes, year of therapy start, sex, ethnicity (major UK groups: White, South Asian, Black, mixed, other ethnicities), social deprivation (index of multiple deprivation quintile), smoking status, the number of current, and ever, prescribed glucose lowering drug classes, baseline HbA_1c_ (closest to treatment start date; range in previous 6 months to +7 days), and other measured clinical features: BMI, eGFR (using the Chronic Kidney Disease Epidemiology Collaboration formula [33]), HDL cholesterol, alanine aminotransferase, albumin, bilirubin, total cholesterol and the mean arterial blood pressure (defined as closest values to treatment start in the previous two years), microvascular complications: nephropathy, neuropathy, retinopathy, and major comorbidities: angina, atherosclerotic cardiovascular disease, atrial fibrillation, cardiac revascularisation, heart failure, hypertension, ischaemic heart disease, myocardial infarction, peripheral arterial disease, stroke, transient ischaemic attack, chronic kidney disease and chronic liver disease.

### Treatment selection model development

#### Model overview

We aimed to build a flexible Bayesian treatment selection model [10] to predict differential glycaemic response that can be readily deployed in clinical practice. The model development process consisted of a first step of propensity score estimation and a second step of developing a treatment selection model using a Bayesian additive regression tree (BART) framework [9, 26]. We aimed to balance predictive accuracy with model parsimony and build the model around a limited number of routinely collected variables, thus facilitating its use in clinical practice. Individuals were excluded from the development and validation sets if they initiated multiple glucose-lowering treatments on the same day; their therapies were initiated less than 61 days since the start of a previous therapy; their baseline HbA_1c_ was <53 mmol/mol; they had a missing baseline HbA_1c_; or they had a missing outcome HbA_1c_ (sFlowchart 1).

The treatment selection model was developed using Bayesian Causal Forests (BCF) [10], a framework specifically designed to estimate heterogeneous treatment effects. The BCF approach places flexible BART priors on the mean functions relating to a prognostic component of the model (representing outcomes in the reference group, in this case, SGLT2i) but also a moderator component that operates over-and-above the prognostic component (in this case, the differential treatment effect of GLP1-RA relative to SGLT2i) [10, 34]. With this method, the shrinkage applied to the differential treatment effects can be adjusted independently of the prognostic effects. We used the bcf [10] and sparseBCF [26] packages in R to fit the model(s) using Markov chain Monte Carlo (MCMC). By default, bcf places stronger regularisation on the moderator side of the model, shrinking the treatment effects towards homogeneity where there is a lack of strong evidence to the contrary. For this model, we are comparing two therapies against each other (rather than a therapy to a control group), so we used the same prior structure for the prognostic and moderator parts of the model. A full description of the model parameters and priors is included in the Supplementary Materials. Currently, the standard BCF software cannot account for missing data [35], so we used a complete case analysis, informed by our previous study showing a limited impact of missing data on predicting CATE in a similar primary care dataset [27].

#### Propensity score estimation

The BCF developers [10] recommend including a propensity score variable in the prognostic component of BCF models to help alleviate regularisation-induced confounding due to prescribing by indication [10]. We used a standard BART model [35] for the propensity score, fitted using MCMC and the bartMachine package in R [35]. All variables extracted initially in the development cohort were used for initial model fitting. However, a subset of the most predictive variables was selected by applying a threshold defined by the proportion of times each predictor was chosen as a spitting rule divided by the total number of splitting rules appearing in the model [35, 36] (sFig. 8). The propensity score model was then refitted with the selected variables. To assess convergence, we monitored the available parameters according to the guidance provided by Kapelner et al. [35] and Gelman-Rubin R̂ values. A description of the model priors is included in the Supplementary Material.

The BART propensity score model converged quickly, so we ran 25,000 iterations with the first 15,000 discarded for burn-in; trace plots are available on request and R̂ < 1.02. To assess the performance of the final model, received operating characteristic (ROC) and precision-recall curves were fitted to both the development and validation cohorts (sFig. 9).

#### Variable selection

Variable selection was deployed to develop a parsimonious final model whilst maintaining predictive accuracy. To do this, we used a two-stage approach, wherein the first stage, we built a sparse BCF model [9] incorporating all candidate predictors. Sparse BCF extends standard BCF by replacing the uniform prior distribution placed over the splitting probabilities of each variable (which means that by default, each variable has the same prior probability of being selected for splitting) with a uniform Dirichlet prior over the splitting probabilities. As the model converges, the posterior distribution for these splitting probabilities induces sparsity by assigning higher weight and, thus, higher variable importance to more predictive covariates. This prior was used in the model’s prognostic and moderator parts [9]. To define the final predictor set, we selected only variables with a posterior mean splitting probability greater than 1/number of variables. This was a subjective choice, but one we found was sufficient to minimise the number of final predictors without meaningfully affecting predictive accuracy. We ran the sparse BCF model for 250,000 iterations, discarding the first 200,000 iterations as burn-in and monitoring convergence using trace plots (available on request) and Gelman-Rubin R̂ values (where all R̂ < 1.01). A description of the model parameters and priors is included in the Supplementary Material.

#### Final model fit

Following variable selection, we fitted a final model using standard BCF without sparsity-inducing priors (since individual-level predictions are not currently possible from the sparse BCF software). The model used 300,000 iterations, discarding the first 200,000 iterations and thinning the remaining iterations by 4, resulting in 25,000 final posterior samples. The propensity score was not included in the final predictor set as it did not meet our threshold for variable selection. As a sensitivity analysis, we refitted the model including the propensity score in the predictor set and compared predictions across the two models (sFig. 11).

#### Variable importance (based on the best linear projection)

Given the known challenge of extracting variable importance from tree-based models, we implemented a pseudo-variable importance measure defined as the proportion of R associated with each variable for predicting the CATE [37]. This was estimated from a linear regression model using all selected variables for the differential part of the model as predictors (with continuous predictors fitted as 3-knot restricted cubic splines) and the predicted CATE as the outcome [38]. To assess how CATE estimates varied across major routine clinical features, we also summarised the marginal distributions of sex, baseline HbA_1c_, eGFR, current age, and BMI across subgroups defined by the degree of predicted glycaemic differences (SGLT2i benefit of 0–3, 3–5 or ≥5 mmol/mol; GLP1-RA benefit of 0–3, 3–5 or ≥5mmol/mol).

### Model validation

Evaluating the accuracy of predicted CATE is a significant challenge since each individual only has outcome data for the treatment they initiated but not for the competing treatment (which they did not take) [21], meaning the treatment effect is not observed at the individual level (it is a so-called counterfactual effect). As such, to validate predicted CATE estimates, we first split validation sets into sub-groups based on their predicted CATE estimates from the BCF model and then compared the average CATE estimate within each sub-group to estimates derived from a set of alternative models fitted to each of the sub-groups in turn. These latter models target the average treatment effect (ATE) within a population of individuals (rather than the conditional average treatment effect [CATE]), with desirable properties justified in the literature [6]. An earlier version of this validation framework is described previously [6]. If the average CATE estimates in each sub-group (from the BCF model) align with the ATE estimates from the alternative models, then this provides evidence that the average treatment effect estimates are consistent across different inference methods within each sub-group. Restricting the ATE estimates for each sub-group allows for simpler comparison ATE models to be used, since the distribution of covariates in each sub-group is expected to be more consistent within each subgroup than for the complete data. For validation, subgroups were defined by decile of predicted CATE in CPRD and, due to lower patient numbers, by quintile in the Tayside & Fife cohort. We then employed three different approaches to estimate the ATE within each subgroup:

#### Regression adjustment (primary approach)

We included all patients and used Bayesian linear regression to estimate the ATE within each subgroup, adjusting for the full covariate set used in the HbA1c treatment selection model (Table 2), with all continuous predictors included as 3-knot restricted cubic splines [39].

#### Propensity score matching

Individuals receiving each drug class within each subgroup were matched by propensity score (the same propensity score used during HbA1c model development), using a caliper distance of 0.05, no replacement and in decreasing order of propensity score values. After defining this restricted patient subset, unadjusted linear regression models were used to estimate the ATE within each subgroup [40].

#### Propensity score matching with adjustment

The linear regression models of approach 2 were refitted using a double robust approach by adjusting for the full covariate set used in the HbA1c treatment selection model (Table 2).

In sensitivity analysis, we further assessed the accuracy of predicted HbA_1c_ treatment effects in those of non-white ethnicity and in those with and without cardiovascular disease. We also evaluated the reproducibility of observed differences in HbA1c response by sex in participants receiving GLP1-RA in the HARMONY clinical trial, the PRIBA prospective study, and Tayside & Fife.

### Secondary outcomes

Specific cohorts were defined to evaluate each secondary outcome to mitigate selection bias and maximise the number of individuals available for analysis (sFlowchart 2). All cohorts required complete predictor data for the HbA1c-based treatment selection model. To evaluate treatment effect heterogeneities, subgroups were defined by the degree of predicted glycaemic differences (SGLT2i benefit of 0–3, 3–5 or ≥5 mmol/mol; GLP1-RA benefit of 0–3, 3–5 or ≥5mmol/mol). As for validation of differences in HbA_1c_ outcomes, three approaches were employed to estimate CATE: regression adjustment (primary approach), propensity score matching, and propensity score matching with predictor variable set adjustment (propensity score model was refitted, following the procedure mentioned before, with the additional inclusion of baseline cardiovascular risk as a predictor (QRISK2 predicted probability of new-onset myocardial infarction or stroke [41]).

For 12-month weight change, we included all patients with a recorded baseline weight (closest value to 2 years prior to treatment initiation) and a valid outcome weight. Treatment effects were estimated using a Bayesian linear regression model with an interaction between the received treatment and the predicted HbA_1c_ treatment benefit subgroup, with adjustment for baseline weight. We included all patients initiating therapy for treatment discontinuation and estimated CATE using Bayesian logistic regression with a treatment-by-HbA_1c_ benefit subgroup interaction. For longer-term outcomes, we included only individuals without the outcome of interest at therapy initiation, thus evaluating only incident events. Patients were followed for up to 5 years using an intention-to-treat approach from the date of therapy initiation until the earliest of: the outcome of interest, the date of GP practice deregistration or death, or the end of the study period. For each outcome, Bayesian Cox proportional hazards models with treatment-by-HbA1c benefit subgroup interactions were fitted with additional adjustment for QRISK2 predicted probability of new-onset myocardial infarction or stroke. A description of priors for each model is included in the Supplementary Material. The models used 10,000 MCMC iterations, discarding the first 5,000 iterations as burn-in for four chains. Convergence was monitored using trace plots and Gelman-Rubin R̂ values. Here all R̂ < 1.04 and trace plots are available on request.

All analyses were conducted using R (version 4.1.2). We followed TRIPOD prediction model reporting guidance (see Supplementary Materials—TRIPOD checklist) [42].

## Acknowledgements

This article is based in part on data from the Clinical Practice Research Datalink obtained under license from the UK Medicines and Healthcare products Regulatory Agency. CPRD data is provided by patients and collected by the NHS as part of their care and support. Approval for CPRD data access and the study protocol was granted by the CPRD Independent Scientific Advisory Committee (eRAP protocol number: 22_002000). This publication is based in part on research using data from GSK that has been made available through secured access. GSK has not contributed to or approved, and is not in any way responsible for, the contents of this publication. The PRIBA study was funded by A National Institute for Health Research (U.K.) Doctoral Research Fellowship (DRF-2010-03-72, AGJ) and supported by the National Institute for Health Research Clinical Research Network. The authors thank the members of the Predicting Response to Incretin Based Agents (PRIBA) study group and all cohort participants. ATH and BMS are supported by the NIHR Exeter Clinical Research Facility; the views expressed are those of the authors and not necessarily those of the NHS, the NIHR or the Department of Health. PC, KGY, JB, TJM and JD are supported by Research England’s Expanding Excellence in England (E3) fund.

## Author contributions

PC, JMD, BS, TJM, ATH, AGJ and ERP conceived and designed the study. PC, with support from JMD, BS and TJM analysed the data and developed the code. KGY, RH and AM helped with curating the CPRD dataset. ATNN, PK, EH and LD helped analyse the Scottish independent dataset. All authors contributed to the writing of the article, provided support for the analysis and interpretation of results, critically revised the article, and approved the final article.

## Code availability statement

All R code used for the analysis is provided at https://github.com/Exeter-Diabetes/CPRD-Pedro-SGLT2vsGLP1.

## Data availability statement

The UK routine clinical data analysed during the current study are available in the CPRD repository (CPRD; https://cprd.com/research-applications), but restrictions apply to the availability of these data, which were used under license for the current study, and so are not publicly available. For re-using these data, an application must be made directly to CPRD. Data from Scotland are anonymized real-world medical records available by request through the Scottish Care Information-Diabetes Collaboration, Tayside & Fife, Scotland unit (https://www.sci-diabetes.scot.nhs.uk/). Clinical trial data are not publicly available, for access an application must be made directly to GSK and www.ClinicalStudyDataRequest.com

---

**Extended Fig. 1:**
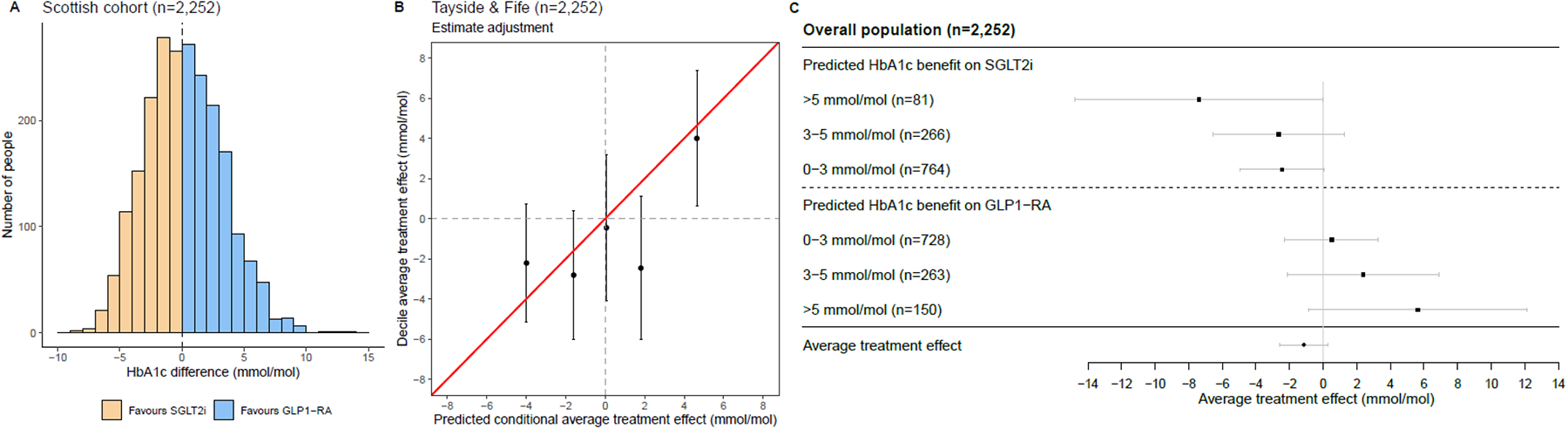
External validation in Tayside & Fife, Scotland. (A) Distribution of conditional average treatment effect (CATE) estimates for SGLT2i vs. GLP1-RA; negative values reflect a predicted glucose-lowering treatment benefit on SGLT2-inhibitors and positive values reflect a predicted treatment benefit on GLP-1 receptor agonists. (B) Calibration between average treatment effects (ATE) and predicted CATE estimates, by quintile of predicted CATE. (C) ATE estimates within subgroups defined by clinically meaningful CATE thresholds (SGLT2i benefit >5, 3-5 and 0-3 mmol/mol, GLP1-RA benefit >5, 3-5 and 0-3 mmol/mol).

**Extended Fig. 2:**
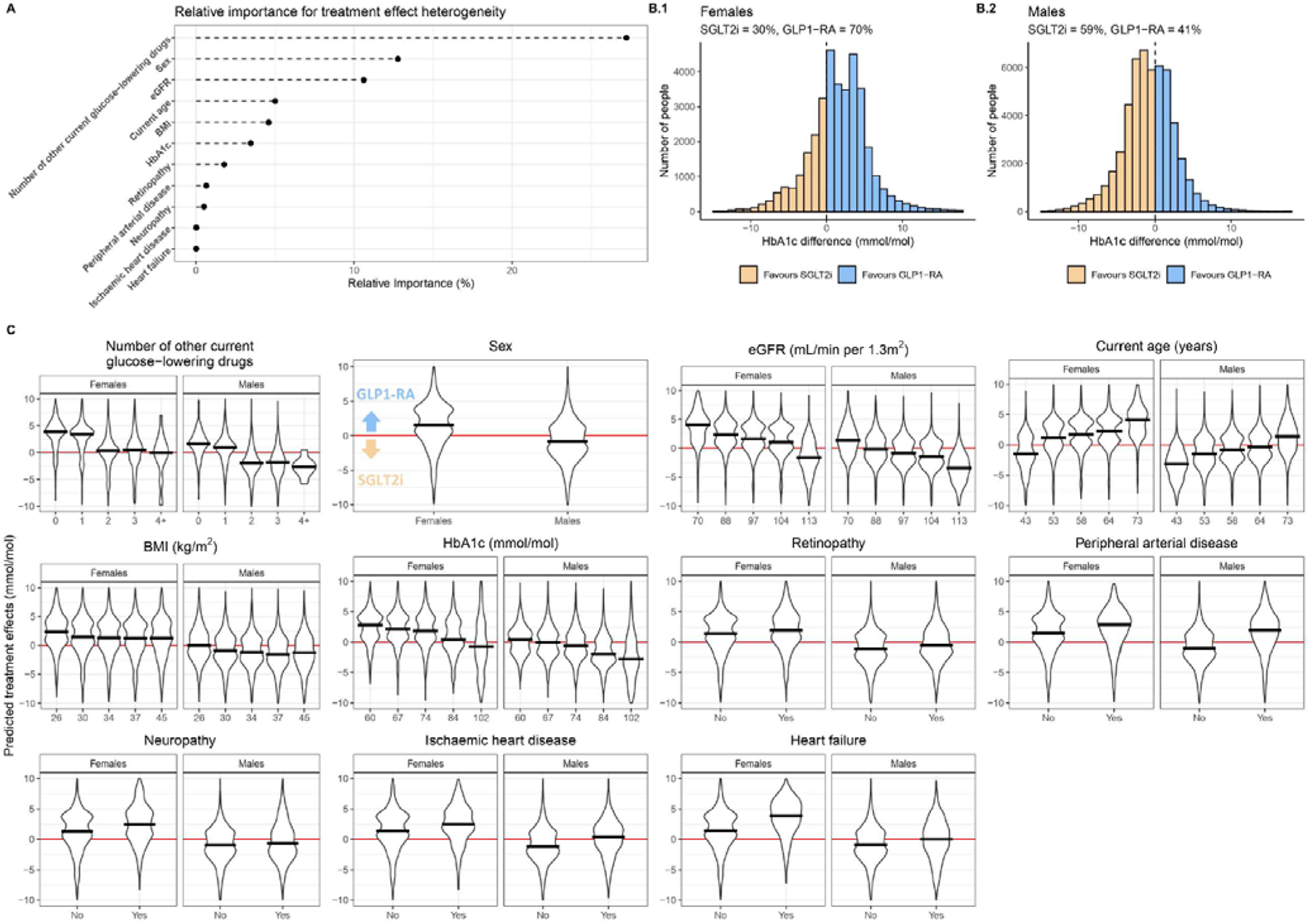
Model interpretability plots. (A) Relative variable importance for clinical features predicting differential treatment effects (best linear projection of BCF model; see Methods). (B) Distribution of conditional average treatment effect (CATE) estimates for SGLT2i vs. GLP1-RA, by sex. (C) Predicted treatment effects for all differential clinical features, with individuals stratified into quintiles for continuous variables, and black lines corresponding to the median of the stratified group. All estimates are for the overall study population, n= 46,394.

**Extended Fig. 3:**
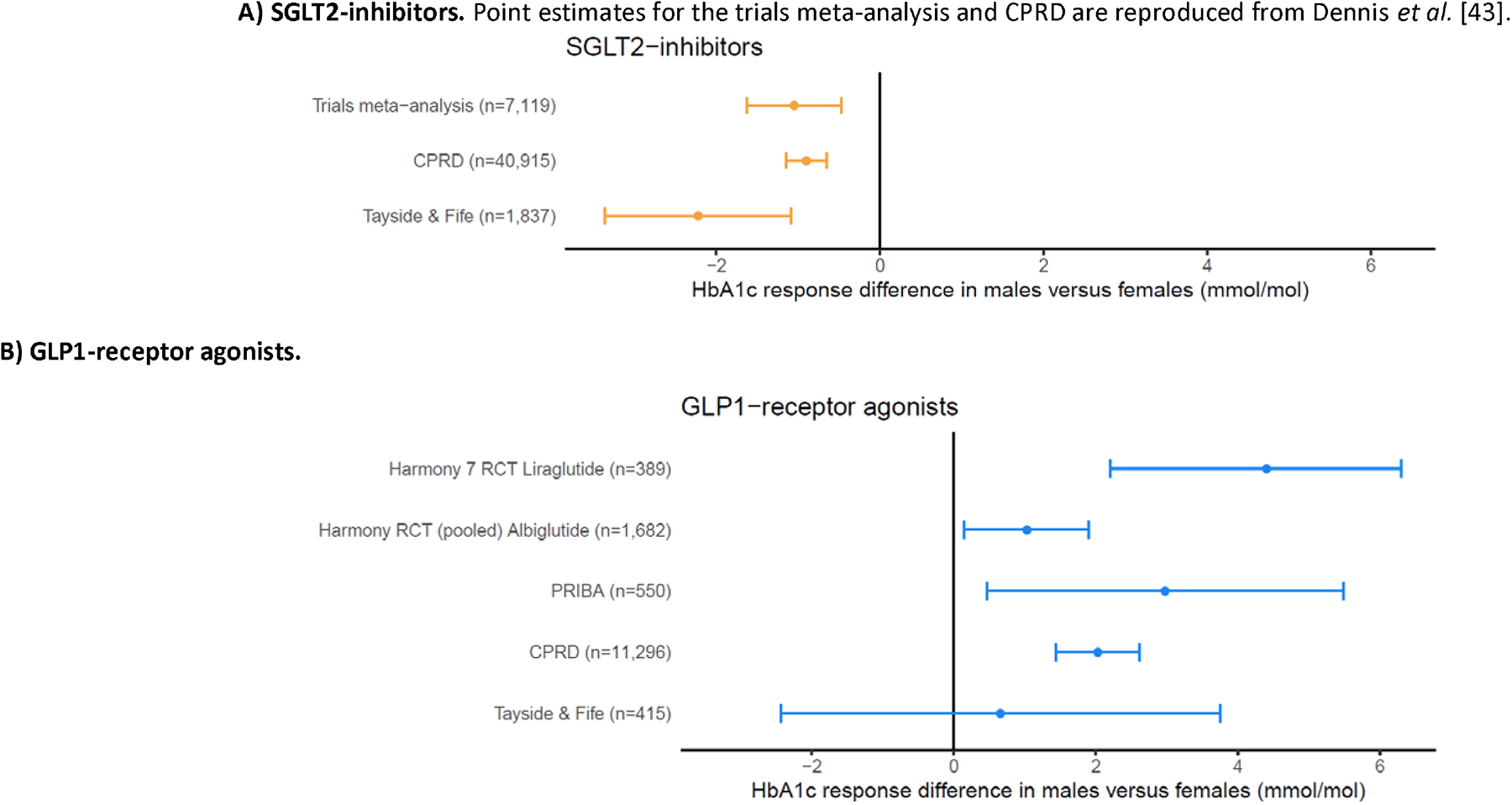
Differences in HbA1c outcome by sex, in randomized clinical trial and observational datasets. All estimates are adjusted for baseline HbA1c. Estimates lower than zero represent a greater HbA1c reduction in males compared to females. Bars represent 95% confidence intervals.

**Extended Fig. 4:**
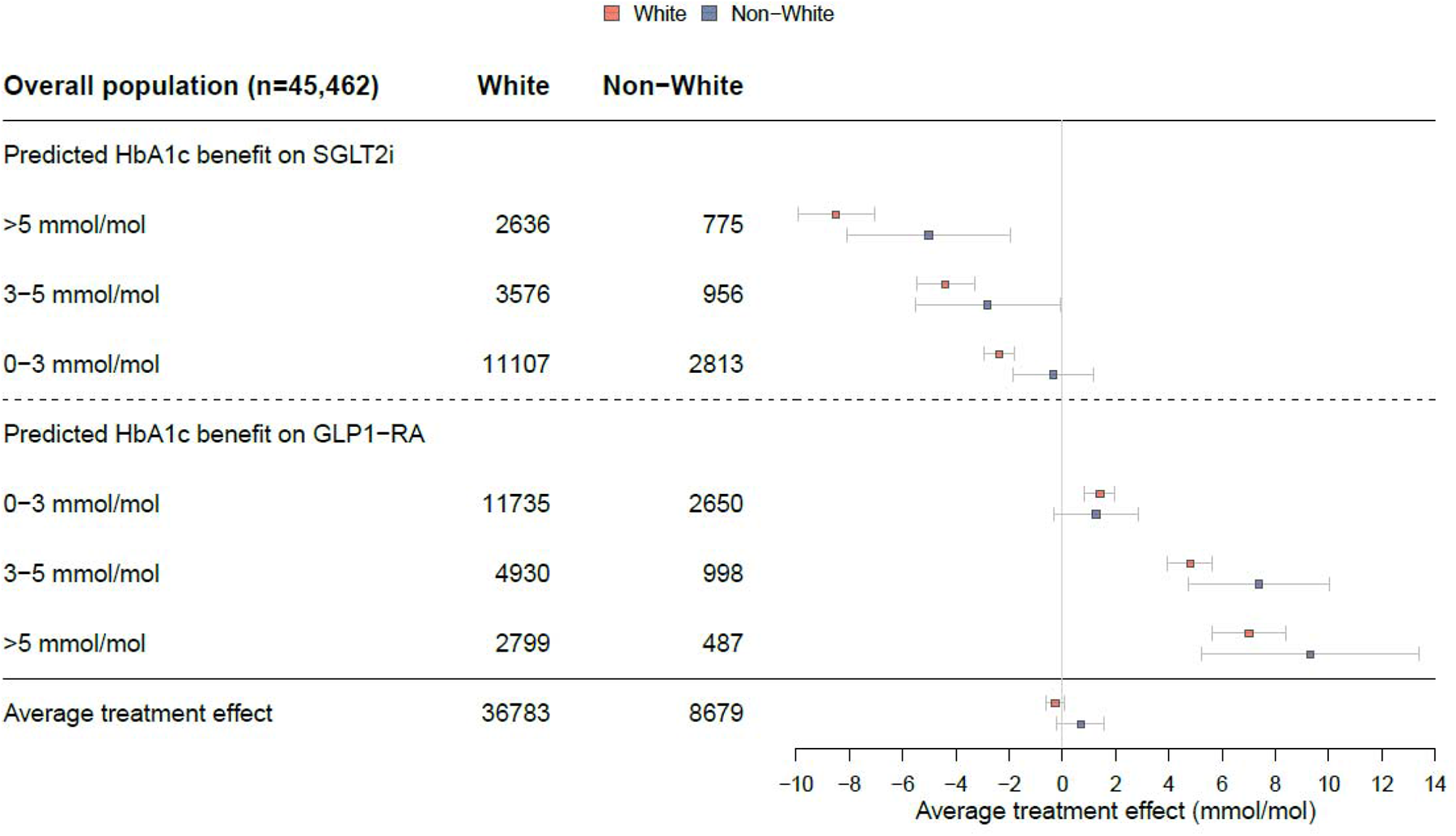
Differential HbA1c treatment effects in individuals of white and non-white ethnicity. Adjusted ATE estimates within subgroups defined by clinically meaningful CATE thresholds (SGLT2i benefit >5, 3-5 and 0-3 mmol/mol, GLP1-RA benefit >5, 3-5 and 0-3 mmol/mol). Negative values reflect a predicted glucose-lowering treatment benefit with SGLT2i, and positive values reflect a predicted treatment benefit with GLP1-RA. The non-white subgroup is a composite of major UK non-white self-reported ethnicity groups: Black, South Asian, Mixed and Other. Individuals without a recorded ethnicity were excluded (n=932)

