## Supplementary Material for "Phenotype-based targeted treatment of SGLT2 inhibitors and GLP-1 receptor agonists in type 2 diabetes"

### Table of Contents

**sFlowchart 1**: CPRD patient flow and inclusion criteria for the development of the treatment selection model. 4

**sFlowchart 2**: CPRD patient flowchart and inclusion criteria for the analysis of additional outcomes. 5

**sTable 1**: Baseline clinical characteristics of patients initiating GLP-1 receptor agonists and SGLT2-inhibitors from the UK Clinical Practice Research Datalink for model derivation and validation cohorts. 6

a) CPRD HbA1c model derivation cohort, n= 31,346. 6

b) CPRD HbA1c model validation cohort, n= 20,865. 9

c) Tayside & Fife (Scotland) routine clinical data, n= 2,252. 11

d) Clinical trials and prospective cohorts. 13

e) CPRD patient subgroups defined by predicted HbA1c benefit >5 mmol/mol with either SGLT2i or GLP-1RA, n= 14,149 (values for Fig. 2). 14

**sTable 2**: Model performance statistics for predicting HbA1c outcome with 95% credible intervals. 16

**sTable 3**: Data underlying Fig. 3, showing differential treatment effects for secondary clinical outcomes across subgroups defined by clinical cut-offs of predicted treatment effects. 17

a) Predicted HbA1c change (n=87,835) 17

b) Predicted weight change (n=41,728), with additional adjustment for baseline weight 17

c) Risk of discontinuation (n=77,741) 17

d) Microvascular complications (n=34,524), with additional adjustment for baseline cardiovascular risk. 18

e) Major adverse cardiovascular events (MACE) (n=52,052), with additional adjustment for baseline cardiovascular risk 18

f) Heart Failure (n=52,052), with additional adjustment for baseline cardiovascular risk 18

g) Developing chronic kidney disease stage 5 or drop of eGFR by 40% (n=52,052), with additional adjustment for baseline cardiovascular risk 19

**sFig. 1**: Variables selected for the prognostic (factors predictive of HbA1c response to SGLT2i therapy) component (A) and the moderator (factors predictive of differential HbA1c response with GLP1-RA compared to SGLT2i therapies) component (B) of the model. 20

**sFig. 2**: Comparison of predicted outcome HbA1c and predicted conditional average treatment effect (CATE) estimates from two Bayesian Causal Forest (BCF) models with and without including propensity scores in the development cohort. 21

**sFig. 3**: Calibration plots of predicted conditional treatment effect (CATE) estimates using propensity score matching. 22

**sFig. 4**: Short-term and long-term clinical outcomes, across subgroups defined by clinical cut-offs of predicted treatment benefit, in propensity score matched cohorts 23

**sFig. 5**: Short-term and long-term clinical outcomes, across subgroups defined by clinical cut-offs of predicted treatment benefit, in propensity score matched cohort with additional covariate adjustment. 24

**sFig. 6**: Relative risk of developing new onset chronic kidney disease (CKD) over 5 years, across subgroups defined by clinical cut-offs of predicted treatment benefit. 25

**sFig. 7**: Evaluation of concordance between estimates of predicted HbA1c outcome with SGLT2i from our model and a recently published SGLT2i-DPP4-inhibitor treatment selection model [1]. 26

**sFig. 8**: Variable selection plot for the propensity score model — identifying the most important predictors. 27

**sFig. 9**: Received operating characteristic (ROC) and precision-recall curves of the propensity score model developed in the derivation cohort and validated in hold-out validation data. 28

**sFig. 10**: Calibration plots of predicted conditional individualised treatment effects in the validation cohort for those with and without cardiovascular disease (CVD). 29

**sFig. 11**: Comparison of predicted outcome HbA1c and predicted conditional average treatment effect (CATE) estimates from a sparse Bayesian Causal Forest (BCF) model and a BCF model with different sets of variables. 30

**TRIPOD Checklist**: Prediction Model Development and Validation 31

Priors of the model. 32

Propensity score model BART 32

Sparse BCF 33

BCF – treatment selection model 34

Additional outcomes 34

References 35

sFlowchart 1: CPRD patient flow and inclusion criteria for the development of the treatment selection model.

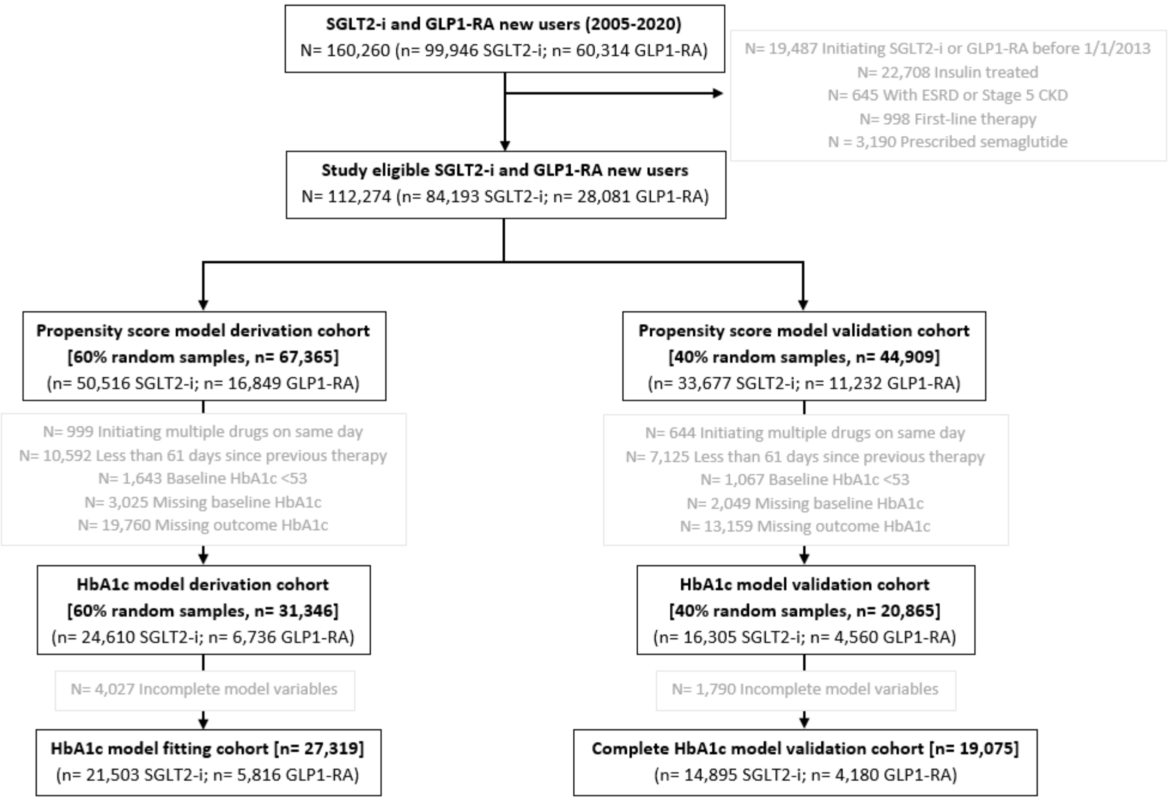


Baseline HbA1c is defined as the closest HbA1c to drug initiation in the previous 6 months. Other biomarkers were defined as the closest measure to drug initiation in the previous 2 years.

sFlowchart 2: CPRD patient flowchart and inclusion criteria for the analysis of additional outcomes.

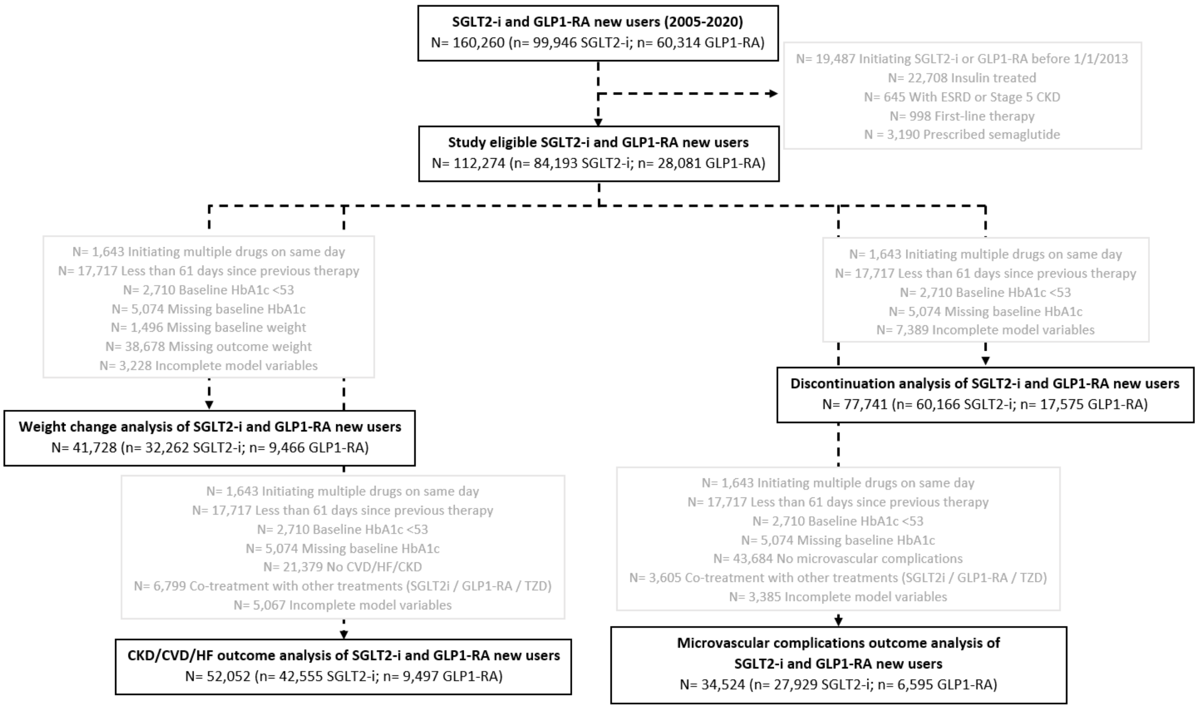

sTable 1: Baseline clinical characteristics of patients initiating GLP-1 receptor agonists and SGLT2-inhibitors from the UK Clinical Practice Research Datalink for model derivation and validation cohorts. **Data are mean [SD] and number (%). Standardised mean difference (SMD). Atherosclerotic cardiovascular disease – composite of myocardial infarction, stroke, ischemic heart disease, peripheral arterial disease and revascularization. *closest values to treatment start in the previous 6 months.**

a) CPRD HbA1c model derivation cohort, n= 31,346.

| | **GLP-1 receptor agonists (n= 6,736)** | | **SGLT2 inhibitors (n= 24,610)** | | **SMD** |
|---|---|---|---|---|---|
| | | Missing (%) | | Missing (%) | |
| Current age, years | 57.8 [10.7] | - | 58.5 [10.4] | - | 0.063 |
| Duration of diabetes, years | 9.3 [6.4] | - | 9.3 [6.3] | - | 0.002 |
| Year of drug start | 2016.4 [10.7] | - | 2017.1 [1.7] | - | 0.389 |
| Sex | | | | | |
| Male | 3,686 (54.7%) | - | 15,264 (62.0%) | - | 0.149 |
| Female | 3,050 (47.3%) | | 9,346 (38.0%) | | |
| Ethnicity | | | | | |
| White | 5,862 (87.0%) | | 18,914 (76.9%) | | |
| South Asian | 461 (6.8%) | 107 (1.6%) | 3,480 (14.1%) | 544 (2.2%) | 0.276 |
| Black | 202 (3.0%) | | 1,042 (4.2%) | | |
| Other | 55 (0.8%) | | 376 (1.5%) | | |
| Mixed | 49 (0.7%) | | 254 (1.0%) | | |
| SGLT2 inhibitor type | | | | | |
| Canagliflozin | | | 4,424 (18.0%) | | |
| Dapagliflozin | - | - | 10,658 (43.3%) | - | |
| Empagliflozin | - | | 9,520 (38.7%) | | |
| Ertugliflozin | - | | 16 (0.1%) | | |
| GLP-1 receptor agonist type | | | | | - |
| Dulaglutide | 2,392 (35.5%) | | - | | |
| Exenatide (short-acting) | 341 (5.1%) | - | - | - | |
| Exenatide (long-acting) | 580 (8.6%) | | - | | |
| Liraglutide | 2,724 (40.4%) | | - | | |
| Lixisenatide | 703 (10.4%) | | - | | |
| Index of multiple deprivation | | | | | |
| 1 (Least deprived) | 1,099 (16.3%) | | 4,207 (17.1%) | | |
| 2 | 1,129 (16.8%) | 2 (<0.1%) | 4,423 (18.0%) | 10 (<0.1%) | 0.059 |
| 3 | 1,345 (20.0%) | | 4,726 (19.2%) | | |
| 4 | 1,433 (21.3%) | | 5,430 (22.1%) | | |
| 5 (Most deprived) | 1,728 (25.7%) | | 5,814 (23.6%) | | |
| Smoking status | | | | | |
| Active | 1,108 (16.4%) | 316 (4.7%) | 3,977 (16.2%) | 993 (4.0%) | 0.061 |
| Ex-smoker | 3,762 (55.8%) | | 13,400 (54.4%) | | |
| Non-smoker | 1,550 (23.0%) | | 6,240 (25.4%) | | |

| | | | | | |
|---|---|---|---|---|---|
| Number of glucose-lowering drug classes ever prescribed | | | | | |
| 2 | 823 (12.2%) | - | 5,815 (23.6%) | - | 0.373 |
| 3 | 1,788 (26.5%) | | 7,734 (31.4%) | | |
| 4 | 2,499 (37.1%) | | 7,151 (29.1%) | | |
| 5+ | 1,626 (24.1%) | | 3,910 (15.9%) | | |
| Number of other current glucose--lowering drugs | | | | | |
| 0 | 278 (4.1%) | | 789 (3.2%) | | |
| 1 | 2,490 (37.0%) | - | 9,898 (40.2%) | - | 0.086 |
| 2 | 3,223 (47.8%) | | 11,173 (45.4%) | | |
| 3 | 720 (10.7%) | | 2,700 (11.0%) | | |
| 4+ | 25 (0.4%) | | 50 (0.2%) | | |
| Background therapy | | | | | |
| Metformin | 6,006 (89.2%) | | 22,465 (91.3%) | | 0.071 |
| Sulfonylurea | 3,273 (48.6%) | | 9,022 (36.7%) | | 0.243 |
| DPP-4 inhibitor | 743 (11.0%) | - | 7,184 (29.2%) | - | 0.465 |
| SGLT2 inhibitor | 857 (12.7%) | | - | | - |
| Thiazolidinedione | 317 (4.7%) | | 616 (2.5%) | | 0.118 |
| GLP-1 receptor agonist | - | | 1,257 (5.1%) | | - |
| Biomarkers | | | | | |
| $HbA_{1c}$, mmol/mol* | 79.2 [15.6] | - | 76.7 [15.6] | - | 0.161 |
| BMI, kg/$m^2$ | 37.7 [7.0] | 122 (1.8%) | 33.6 [6.7] | 781 (3.2%) | 0.589 |
| eGFR, mL/min per 1.3 $m^2$ | 92.6 [18.7] | 6 (0.1%) | 94.9 [14.8] | 19 (0.1%) | 0.139 |
| HDL, cholesterol, mmol/L | 1.1 [0.3] | 328 (4.9%) | 1.1 [0.3] | 774 (3.1%) | 0.084 |
| Alanine transaminase, IU/L | 35.8 [20.4] | 418 (6.2%) | 35.2 [20.2] | 1,395 (5.7%) | 0.033 |
| Albumin, g/L | 41.6 [3.9] | 328 (4.9%) | 42.1 [3.9] | 1,010 (4.1%) | 0.124 |
| Bilirubin, µmol/L | 9.1 [4.7] | 265 (3.9%) | 9.5 [4.9] | 914 (3.7%) | 0.087 |
| Total cholesterol, mmol/L | 4.3 [1.1] | 17 (0.3%) | 4.2 [1.1] | 28 (0.1%) | 0.071 |
| Mean arterial blood pressure, mm Hg | 96.1 [8.8] | 12 (0.2%) | 96.0 [8.8] | 36 (0.1%) | 0.010 |
| Microvascular complications | | | | | |
| Nephropathy | 173 (2.6%) | - | 461 (1.9%) | - | 0.047 |
| Neuropathy | 1,828 (27.1%) | - | 5,825 (23.7%) | - | 0.080 |
| Retinopathy | 2,438 (36.2%) | - | 9,332 (37.9%) | - | 0.036 |
| Cardiovascular conditions | | | | | |
| Angina | 768 (11.4%) | - | 2,308 (9.4%) | - | 0.066 |
| Atherosclerotic cardiovascular disease | 1,420 (21.1%) | - | 4,551 (18.5%) | - | 0.065 |
| Atrial fibrillation | 407 (6.0%) | - | 1,094 (4.4%) | - | 0.072 |
| Cardiac revascularisation | 420 (6.2%) | - | 1,520 (6.2%) | - | 0.002 |
| Heart failure | 380 (5.6%) | - | 913 (3.7%) | - | 0.092 |
| Hypertension | 4,057 (60.2%) | - | 13,778 (56.0%) | - | 0.086 |
| Ischaemic heart disease | 972 (14.4%) | - | 3,125 (12.7%) | - | 0.051 |
| Myocardial infarction | 466 (6.9%) | - | 1,546 (6.3%) | - | 0.026 |
| Peripheral arterial disease | 347 (5.2%) | - | 1,041 (4.2%) | - | 0.044 |
| Stroke | 280 (4.2%) | - | 914 (3.7%) | - | 0.023 |
| Transient ischaemic attack | 152 (2.3%) | - | 557 (2.3%) | - | <0.001 |

| | | | | | |
|---|---|---|---|---|---|
| Other conditions | | | | | |
| Chronic kidney disease | 561 (8.3%) | - | 731 (3.0%) | - | 0.234 |
| Chronic liver disease | 907 (13.5%) | - | 2,922 (11.9%) | - | 0.048 |
| QRISK2 10-year score | 23.5 [13.4] | 308 (4.6%) | 23.2 [12.8] | 1,611 (6.5%) | 0.021 |
| $HbA_{1c}$ outcome | | | | | |
| $HbA_{1c}$, mmol/mol | 67.0 [18.2] | - | 64.5 [15.0] | - | 0.152 |
| Month of $HbA_{1c}$ measure | 9.0 [3.5] | - | 9.2 [3.5] | - | 0.067 |

b) CPRD HbA1c model validation cohort, n= 20,865.

| | **GLP-1 receptor agonists (n= 4,560)** | | **SGLT2 inhibitors (n= 16,305)** | | **SMD** |
|---|---|---|---|---|---|
| | | Missing (%) | | Missing (%) | |
| Current age, years | 58.2 [10.8] | - | 58.3 [10.3] | - | 0.009 |
| Duration of diabetes, years | 9.3 [6.1] | - | 9.2 [6.3] | - | 0.025 |
| Year of drug start | 2016.4 [2.1] | - | 2017.1 [1.7] | - | 0.378 |
| Sex | | | | | |
| Male | 2,460 (53.9%) | - | 10,298 (63.2%) | - | 0.188 |
| Female | 2,100 (46.1%) | | 6,007 (36.8%) | | |
| Ethnicity | | | | | |
| White | 3,995 (87.6%) | | 12,596 (77.3%) | | |
| South Asian | 293 (6.4%) | 60 | 2,267 (13.9%) | 389 | 0.289 |
| Black | 133 (2.9%) | (1.3%) | 653 (4.0%) | (2.4%) | |
| Other | 38 (0.8%) | | 249 (1.5%) | | |
| Mixed | 41 (0.9%) | | 151 (0.9%) | | |
| SGLT2 inhibitor type | | | | | |
| Canagliflozin | - | | 2,921 (17.9%) | | |
| Dapagliflozin | - | - | 7,182 (44.0%) | - | |
| Empagliflozin | - | | 6,190 (38.0%) | | |
| Ertugliflozin | - | | 13 (0.1%) | | |
| GLP-1 receptor agonist type | | | | | - |
| Dulaglutide | 1,621 (35.5%) | | - | | |
| Exenatide (short-acting) | 228 (5.0%) | | - | | |
| Exenatide (long-acting) | 380 (8.3%) | - | - | - | |
| Liraglutide | 1,884 (41.3%) | | - | | |
| Lixisenatide | 449 (9.8%) | | - | | |
| Index of multiple deprivation | | | | | |
| 1 (Least deprived) | 784 (17.2%) | | 2,810 (17.2%) | | |
| 2 | 823 (18.0%) | 3 | 2,905 (17.8%) | 8 | 0.043 |
| 3 | 878 (19.3%) | (0.1%) | 3,197 (19.6%) | (<0.1%) | |
| 4 | 949 (20.8%) | | 3,609 (22.1%) | | |
| 5 (Most deprived) | 1,123 (24.6%) | | 3,776 (23.2%) | | |
| Smoking status | | | | | |
| Active | 760 (16.7%) | 204 | 2,595 (15.9%) | 670 | 0.055 |
| Ex-smoker | 2,539 (55.7%) | (4.5%) | 8,890 (54.5%) | (4.1%) | |
| Non-smoker | 1,057 (23.2%) | | 4,150 (25.5%) | | |
| Number of glucose-lowering drug classes ever prescribed | | | | | |
| 2 | 564 (12.4%) | | 3,920 (24.0%) | | |
| 3 | 1,232 (27.0%) | - | 5,164 (31.7%) | - | 0.378 |
| 4 | 1,652 (36.2%) | | 4,663 (28.6%) | | |
| 5+ | 1,112 (24.4%) | | 2,558 (15.7%) | | |

| | | | | | |
|---|---|---|---|---|---|
| Number of other current glucose--lowering drugs | | | | | |
| 0 | 199 (4.4%) | | 561 (3.4%) | | |
| 1 | 1,682 (36.9%) | - | 6,644 (40.7%) | - | 0.098 |
| 2 | 2,141 (47.0%) | | 7,364 (45.2%) | | |
| 3 | 512 (11.2%) | | 1,692 (10.4%) | | |
| 4+ | 26 (0.6%) | | 44 (0.3%) | | |
| Background therapy | | | | | |
| Metformin | 4,048 (88.8%) | | 14,961 (91.8%) | | 0.101 |
| Sulfonylurea | 2,206 (48.4%) | | 5,851 (35.9%) | | 0.255 |
| DPP-4 inhibitor | 505 (11.1%) | - | 4,584 (28.1%) | - | 0.440 |
| SGLT2 inhibitor | 651 (14.3%) | | - | | - |
| Thiazolidinedione | 195 (4.3%) | | 418 (2.6%) | | 0.094 |
| GLP-1 receptor agonist | - | | 810 (2.6%) | | - |
| Biomarkers | | | | | |
| $HbA_{1c}$, mmol/mol* | 78.8 [15.5] | - | 76.8 [15.3] | - | 0.138 |
| BMI, kg/$m^2$ | 37.4 [6.9] | 104 (2.3%) | 33.7 [6.6] | 508 (3.1%) | 0.549 |
| eGFR, mL/min per 1.3 $m^2$ | 91.8 [19.5] | 1 (<0.1%) | 95.2 [14.7] | 14 (0.1%) | 0.196 |
| HDL, cholesterol, mmol/L | 1.1 [0.3] | 192 (4.2%) | 1.1 [0.3] | 560 (3.4%) | 0.074 |
| Alanine transaminase, IU/L | 35.9 [21.0] | 283 (6.2%) | 35.5 [20.5] | 924 (5.7%) | 0.018 |
| Albumin, g/L | 41.7 [3.9] | 197 (4.3%) | 42.1 [3.9] | 725 (4.4%) | 0.111 |
| Bilirubin, µmol/L | 9.1 [4.4] | 168 (3.7%) | 9.6 [5.0] | 640 (3.9%) | 0.101 |
| Total cholesterol, mmol/L | 4.3 [1.1] | 9 (0.2%) | 4.2 [1.1] | 25 (0.2%) | 0.087 |
| Mean arterial blood pressure, mm Hg | 96.1 [9.0] | 5 (0.1%) | 96.2 [8.7] | 37 (0.2%) | 0.011 |
| Microvascular complications | | | | | |
| Nephropathy | 112 (2.5%) | - | 328 (2.0%) | - | 0.030 |
| Neuropathy | 1,256 (27.5%) | - | 3,780 (23.2%) | - | 0.100 |
| Retinopathy | 1,706 (37.4%) | - | 6,097 (37.4%) | - | <0.001 |
| Cardiovascular conditions | | | | | |
| Angina | 540 (11.8%) | - | 1,446 (8.9%) | - | 0.098 |
| Atherosclerotic cardiovascular disease | 1,086 (23.8%) | - | 2,979 (18.3%) | - | 0.136 |
| Atrial fibrillation | 286 (6.3%) | - | 726 (4.5%) | - | 0.081 |
| Cardiac revascularisation | 319 (7.0%) | - | 1,054 (6.5%) | - | 0.021 |
| Heart failure | 249 (5.5%) | - | 600 (3.7%) | - | 0.085 |
| Hypertension | 2,774 (60.8%) | - | 9,089 (55.7%) | - | 0.103 |
| Ischaemic heart disease | 703 (15.4%) | - | 1,979 (12.1%) | - | 0.095 |
| Myocardial infarction | 319 (7.0%) | - | 998 (6.1%) | - | 0.035 |
| Peripheral arterial disease | 291 (6.4%) | - | 675 (4.1%) | - | 0.101 |
| Stroke | 238 (5.2%) | - | 617 (3.8%) | - | 0.069 |
| Transient ischaemic attack | 148 (3.2%) | - | 377 (2.3%) | - | 0.057 |
| Other conditions | | | | | |
| Chronic kidney disease | 460 (10.1%) | - | 447 (2.7%) | - | 0.303 |
| Chronic liver disease | 603 (13.2%) | - | 1,917 (11.8%) | - | 0.044 |
| QRISK2 10-year score | 24.0 [13.3] | 224 (4.9%) | 23.1 [12.8] | 1,056 (6.5%) | 0.067 |
| $HbA_{1c}$ outcome | | | | | |
| $HbA_{1c}$, mmol/mol | 67.0 [17.8] | - | 64.4 [14.7] | - | 0.161 |
| Month of $HbA_{1c}$ measure | 8.9 [3.5] | - | 9.2 [3.5] | - | 0.089 |

c) Tayside & Fife (Scotland) routine clinical data, n= 2,252.

| | **GLP-1 receptor agonists (n= 415)** | | **SGLT2 inhibitors (n= 1,837)** | |
|---|---|---|---|---|
| | | Missing (%) | | Missing (%) |
| Current age, years | 58.7 [9.1] | - | 61.5 [9.7] | - |
| Duration of diabetes, years | 7.5 [4.2] | - | 7.4 [4.6] | - |
| Year of drug start | 2,013.3 [3.2] | - | 2,017 [1.4] | - |
| Sex | | | | |
| Male | 226 (54.5%) | - | 1,155 (62.9%) | - |
| Female | 189 (45.5%) | | 682 (37.1%) | |
| Ethnicity | | | | |
| White | 415 (100%) | | 1,837 (100%) | |
| South Asian | - | - | - | - |
| Black | - | | - | |
| Other | - | | - | |
| Mixed | - | | - | |
| Index of multiple deprivation | | | | |
| 1 (Least deprived) | 93 (23.2%) | | 375 (21.1%) | |
| 2 | 91 (22.7%) | 14 | 407 (22.9%) | 63 |
| 3 | 74 (18.5%) | (3.4%) | 355 (20.0%) | (3.4%) |
| 4 | 73 (18.2%) | | 348 (19.6%) | |
| 5 (Most deprived) | 70 (17.5%) | | 289 (16.3%) | |
| Smoking status | | | | |
| Active | 296 (71.3%) | - | 1,258 (68.5%) | - |
| Ex-smoker | - | | - | |
| Non-smoker | 119 (28.7%) | | 579 (31.5%) | |
| Number of glucose-lowering drug classes ever prescribed | | | | |
| 2 | 43 (10.4%) | - | 580 (31.6%) | - |
| 3 | 125 (30.1%) | | 612 (33.3%) | |
| 4 | 148 (35.7%) | | 412 (22.4%) | |
| 5+ | 99 (23.9%) | | 233 (12.7%) | |
| Number of other current glucose--lowering drugs | | - | | - |
| 0 / 1 | 109 (26.3%) | | 861 (46.9%) | |
| 2 + | 306 (73.7%) | | 976 (53.1%) | |
| Biomarkers | | | | |
| $HbA_{1c}$, mmol/mol* | 82.8 [16.8] | - | 76.9 [14.3] | - |
| BMI, kg/m$^2$ | 38.8 [7.4] | - | 34.4 [6.6] | - |
| eGFR, mL/min per 1.3 m$^2$ | 93.2 [19.1] | - | 92.5 [16.0] | - |
| HDL, cholesterol, mmol/L | 1.1 [0.3] | - | 1.1 [0.3] | - |
| Alanine transaminase, IU/L | 38.1 [21.3] | - | 39.3 [23.2] | - |
| Albumin, g/L | 40.9 [4.1] | - | 39.6 [3.8] | - |
| Bilirubin, µmol/L | 8.7 [4.1] | - | 9.7 [4.6] | - |
| Total cholesterol, mmol/L | 4.4 [1.2] | 1 (0.2%) | 4.3 [1.1] | 3 (0.2%) |
| Mean arterial blood pressure, mm Hg | 100.8 [11.6] | 3 (0.7%) | 97.6 [9.7] | 8 (0.4%) |
| Microvascular complications | | | | |
| Retinopathy | 193 (46.5%) | - | 779 (42.4%) | - |

| | | | | |
|---|---|---|---|---|
| Cardiovascular conditions | | | | |
| Atrial fibrillation | 23 (5.5%) | - | 85 (4.6%) | - |
| Heart failure | 19 (4.6%) | - | 41 (2.2%) | - |
| Myocardial infarction | 24 (5.8%) | - | 141 (7.7%) | - |
| Peripheral arterial disease | 2 (0.5%) | - | 11 (0.6%) | - |
| Stroke | 7 (1.7%) | - | 40 (2.2%) | - |
| Transient ischaemic attack | 2 (0.5%) | - | 13 (0.7%) | - |
| $HbA_{1c}$ outcome | | | | |
| $HbA_{1c}$, mmol/mol | 68.2 [17.7] | - | 63.5 [13.8] | - |
| Month of $HbA_{1c}$ measure | 9.5 [3.2] | - | 9.6 [3.3] | - |

d) Clinical trials and prospective cohorts.

| | HARMONY 7 RCT: Liraglutide (n= 389) | HARMONY 7 RCT: Albiglutide (n= 1,682) | PRIBA (n= 550) |
|---|---|---|---|
| Current age, years | 55.8 [10.0] | 56.4 [10.0] | 55.8 [10.3] |
| Duration of diabetes, years | 8.3 [5.6] | 8.1 [6.4] | 10.0 [6.5] |
| Sex | | | |
| Male | 203 (52.2%) | 873 (52.0%) | 299 (54.4%) |
| Female | 186 (47.8%) | 809 (48.0%) | 251 (45.6%) |
| Ethnicity | | | |
| White | 275 (70.7%) | 1,127 (67.0%) | 299 (54.4%) |
| Non-white | 114 (29.3%) | 555 (33.0%) | 251 (45.6%) |
| Number of other current glucose--lowering drugs | | | |
| 0 / 1 | 341 (87.6%) | 1,476 (87.7%) | 257 (46.7%) |
| 2 + | 48 (12.4%) | 206 (12.2%) | 293 (53.3%) |
| Biomarkers | | | |
| $HbA_{1c}$, mmol/mol | 66.0 [14.3] | 65.9 [9.9] | 82.6 [17.6] |
| BMI, kg/$m^2$ | 32.0 [5.9] | 32.7 [5.7] | 39.6 [7.5] |
| eGFR, mL/min per 1.3 $m^2$ | 95.3 [16.6] | 86.5 [20.4] | 92.4 [26.6] |
| HDL, cholesterol, mmol/L | 1.2 [0.3] | 1.2 [0.3] | 1.1 [0.6] |
| Alanine transaminase, IU/L | 27.7 [13.6] | 26.4 [15.7] | 34.4 [19.3] |
| Albumin, g/L | 25.9 [37.2] | 28.8 [37.5] | 41.6 [14.8] |
| Bilirubin, µmol/L | 9.6 [4.3] | 9.5 [4.2] | 9.5 [6.4] |
| Total cholesterol, mmol/L | 4.5 [1.0] | 4.7 [1.1] | 4.4 [1.2] |
| $HbA_{1c}$ outcome | | | |
| $HbA_{1c}$, mmol/mol | 54.0 [12.8] | 55.0 [13.9] | 68.0 [16.8] |

e) CPRD patient subgroups defined by predicted HbA1c benefit >5 mmol/mol with either SGLT2i or GLP-1RA, n= 14,149 (values for Fig. 2).

| | **Predicted benefit on GLP1-RA >5 mmol/mol (n= 3,319)** | | **Predicted benefit on SGLT2i >5 mmol/mol (n= 3,485)** | | **SMD** |
|---|---|---|---|---|---|
| | | Missing (%) | | Missing (%) | |
| Current age, years | 69.4 [9.2] | - | 48.8 [10.1] | - | 2.129 |
| Duration of diabetes, years | 11.5 [7.4] | - | 8.1 [5.4] | - | 0.525 |
| Year of drug start | 2,017.3 [1.8] | - | 2,016.7 [1.9] | - | 0.286 |
| Therapy taken | | | | | |
| GLP1-RA | 956 (28.8%) | - | 904 (25.9%) | - | 0.064 |
| SGLT2i | 2,363 (71.2%) | | 2,581 (74.1%) | | |
| Sex | | | | | |
| Male | 906 (27.3%) | - | 2,550 (73.2%) | - | 1.033 |
| Female | 2,413 (72.7%) | | 935 (26.8%) | | |
| Ethnicity | | | | | |
| White | 2,799 (84.3%) | | 2,636 (75.6%) | | |
| South Asian | 305 (9.2%) | 33 | 555 (15.9%) | 74 | 0.249 |
| Black | 133 (4.0%) | (1.0%) | 127 (3.6%) | (2.1%) | |
| Other | 28 (0.8%) | | 56 (1.6%) | | |
| Mixed | 21 (0.6%) | | 37 (1.1%) | | |
| SGLT2 inhibitor type | | | | | |
| Canagliflozin | 490 (14.8%) | - | 453 (13.0%) | - | |
| Dapagliflozin | 845 (25.5%) | - | 1,217 (34.9%) | - | |
| Empagliflozin | 1,029 (30.8%) | - | 912 (26.2%) | - | |
| Ertugliflozin | 0 (0%) | - | 0 (0%) | - | |
| GLP-1 receptor agonist type | | | | | - |
| Dulaglutide | 389 (11.7%) | - | 285 (8.2%) | - | |
| Exenatide (short-acting) | 35 (1.1%) | - | 57 (1.6%) | - | |
| Exenatide (long-acting) | 71 (2.1%) | - | 81 (2.3%) | - | |
| Liraglutide | 384 (11.6%) | - | 388 (11.1%) | - | |
| Lixisenatide | 77 (2.3%) | - | 93 (2.7%) | - | |
| Index of multiple deprivation | | | | | |
| 1 (Least deprived) | 671 (20.2%) | | 473 (13.6%) | | |
| 2 | 608 (18.3%) | 2 | 525 (15.1%) | 1 | 0.249 |
| 3 | 668 (20.1%) | (0.1%) | 665 (19.1%) | (<0.1%) | |
| 4 | 680 (20.5%) | | 841 (24.1%) | | |
| 5 (Most deprived) | 690 (20.8%) | | 980 (28.1%) | | |
| Smoking status | | | | | |
| Active | 346 (10.4%) | 147 | 779 (22.4%) | 140 | 0.348 |
| Ex-smoker | 2,019 (60.8%) | (4.4%) | 1,677 (48.1%) | (4.0%) | |
| Non-smoker | 807 (24.3%) | | 889 (25.5%) | | |
| Number of glucose-lowering drug classes ever prescribed | | | | | |
| 2 | 792 (23.9%) | - | 276 (7.9%) | - | 0.450 |
| 3 | 957 (28.8%) | | 1,144 (32.8%) | | |
| 4 | 966 (29.1%) | | 1,307 (37.5%) | | |
| 5+ | 604 (18.2%) | | 758 (21.8%) | | |

| | | | | | |
|---|---|---|---|---|---|
| Number of other current glucose--lowering drugs | | | | | |
| 0 | 385 (11.6%) | | 25 (0.7%) | | |
| 1 | 2,027 (61.1%) | - | 467 (13.4%) | - | 1.477 |
| 2 | 731 (22.0%) | | 2,346 (67.3%) | | |
| 3 | 175 (5.3%) | | 623 (17.9%) | | |
| 4+ | 1 (<0.1%) | | 24 (0.7%) | | |
| Background therapy | | | | | |
| Metformin | 2,424 (73.0%) | - | 3,362 (96.5%) | - | 0.690 |
| Sulfonylurea | 910 (27.4%) | - | 2,046 (58.7%) | - | 0.666 |
| DPP-4 inhibitor | 556 (16.8%) | - | 1,133 (32.5%) | - | 0.372 |
| SGLT2 inhibitor | 2,393 (72.1%) | - | 2,731 (78.4%) | - | 0.146 |
| Thiazolidinedione | 57 (1.7%) | - | 142 (4.1%) | - | 0.141 |
| GLP-1 receptor agonist | 997 (30.0%) | - | 1,195 (34.3%) | - | 0.091 |
| Biomarkers | | | | | |
| $HbA_{1c}$, mmol/mol* | 78.6 [20.6] | - | 94.5 [15.5] | - | 0.871 |
| BMI, kg/$m^2$ | 33.6 [8.9] | - | 36.0 [6.2] | - | 0.310 |
| eGFR, mL/min per 1.3 $m^2$ | 74.9 [17.9] | - | 108.4 [13.6] | - | 2.106 |
| HDL, cholesterol, mmol/L | 1.2 [0.3] | 36 (1.1%) | 1.0 [0.3] | 52 (1.5%) | 0.661 |
| Alanine transaminase, IU/L | 27.4 [15.4] | - | 40.8 [23.6] | - | 0.672 |
| Albumin, g/L | 40.9 [4.0] | 23 (0.7%) | 42.0 [3.9] | 22 (0.6%) | 0.285 |
| Bilirubin, µmol/L | 8.8 [4.5] | 8 (0.2%) | 9.6 [5.1] | 13 (0.4%) | 0.179 |
| Total cholesterol, mmol/L | 4.3 [1.1] | 1 (<0.1%) | 4.4 [1.1] | 1 (<0.1%) | 0.089 |
| Mean arterial blood pressure, mm Hg | 94.3 [8.9] | 0 (0%) | 97.1 [8.9] | 4 (0.1%) | 0.315 |
| Microvascular complications | | | | | |
| Nephropathy | 90 (2.7%) | - | 90 (2.6%) | - | 0.008 |
| Neuropathy | 1,536 (46.3%) | - | 531 (15.2%) | - | 0.714 |
| Retinopathy | 1,837 (55.3%) | - | 1,004 (28.8%) | - | 0.558 |
| Cardiovascular conditions | | | | | |
| Angina | 472 (14.2%) | - | 226 (6.5%) | - | 0.256 |
| Atherosclerotic cardiovascular disease | 974 (29.3%) | - | 442 (12.7%) | - | 0.418 |
| Atrial fibrillation | 341 (10.3%) | - | 119 (3.4%) | - | 0.274 |
| Cardiac revascularisation | 242 (7.3%) | - | 128 (3.7%) | - | 0.159 |
| Heart failure | 293 (8.8%) | - | 147 (4.2%) | - | 0.188 |
| Hypertension | 2,317 (69.8%) | - | 1,601 (45.9%) | - | 0.498 |
| Ischaemic heart disease | 580 (17.5%) | - | 297 (8.5%) | - | 0.269 |
| Myocardial infarction | 256 (7.7%) | - | 146 (4.2%) | - | 0.149 |
| Peripheral arterial disease | 332 (10.0%) | - | 95 (2.7%) | - | 0.301 |
| Stroke | 254 (7.7%) | - | 103 (3.0%) | - | 0.211 |
| Transient ischaemic attack | 175 (5.3%) | - | 49 (1.4%) | - | 0.216 |
| Other conditions | | | | | |
| Chronic kidney disease | 832 (25.1%) | - | 33 (0.9%) | - | 0.768 |
| Chronic liver disease | 394 (11.9%) | - | 493 (14.1%) | - | 0.068 |
| QRISK2 10-year score | 32.0 [14.5] | 196 (5.9%) | 18.3 [11.8] | 156 (4.5%) | 1.040 |
| $HbA_{1c}$ outcome | | | | | |
| $HbA_{1c}$, mmol/mol | 67.3 [18.9] | - | 73.5 [18.6] | - | 0.326 |
| Month of $HbA_{1c}$ measure | 8.7 [3.6] | | 9.2 [3.5] | | 0.131 |

sTable 2: Model performance statistics for predicting HbA1c outcome with 95% credible intervals. $R^2$ and root mean square error (RMSE) were derived from 25,000 iterations post-convergence of the Hba1c treatment selection model.

| | Internal validation (development data: n=27,319) | Internal validation (hold-back data: n=19,075) |
|---|---|---|
| $R^2$ | 0.28 (0.28, 0.29) | 0.26 (0.26, 0.27) |
| RMSE (mmol/mol) | 13.4 (13.3, 13.4) | 13.4 (13.3, 13.5) |

sTable 3: Data underlying Fig. 3, showing differential treatment effects for secondary clinical outcomes across subgroups defined by clinical cut-offs of predicted treatment effects.
Estimated values are adjusted for the clinical features used in the treatment selection model (to improve precision and control for potential differences in covariate balance within subgroups).

a) Predicted HbA1c change (n=87,835)

| Predicted HbA1c benefit | Predicted HbA1c change (mmol/mol) | | |
|---|---|---|---|
| | N patients | Predicted HbA1c change on SGLT2i (95% CI) | Predicted HbA1c change on GLP1-RA (95% CI) |
| Overall | 87,835 | -12.0 (-12.1, -11.9) | -12.0 (-12.3, -11.8) |
| Subgroup | | | |
| SGLT2i benefit by ≥5 mmol/mol | 6,856 | -23.3 (-24.0, -22.6) | -18.4 (-19.3, -17.6) |
| SGLT2i benefit by 3-5 mmol/mol | 8,643 | -17.2 (-17.8, -16.7) | -12.9 (-13.7, -12.2) |
| SGLT2i benefit by 0-3 mmol/mol | 26,088 | -12.6 (-12.9, -12.3) | -10.3 (-10.8, -9.9) |
| GLP1-RA benefit by 0-3 mmol/mol | 27,415 | -9.6 (-9.9, -9.4) | -10.8 (-11.3, -10.3) |
| GLP1-RA benefit by 3-5 mmol/mol | 11,540 | -7.4 (-7.8, -7.0) | -12.0 (-12.7, -11.3) |
| GLP1-RA benefit by ≥5 mmol/mol | 7,293 | -9.0 (-9.7, -8.2) | -15.7 (-16.6, -14.8) |

b) Predicted weight change (n=41,728), with additional adjustment for baseline weight

| Predicted HbA1c benefit | Predicted weight change (mmol/mol) | | |
|---|---|---|---|
| | N patients | Predicted weight change on SGLT2i (95% CI) | Predicted weight change on GLP1-RA (95% CI) |
| Overall | 41,728 | -4.20 (-4.3, -4.2) | -2.8 (-2.9, -2.7) |
| Subgroup | | | |
| SGLT2i benefit by ≥5 mmol/mol | 3,152 | -3.7 (-3.9, -3.4) | -2.4 (-2.8, -2.0) |
| SGLT2i benefit by 3-5 mmol/mol | 4,196 | -3.9 (-4.1, -3.7) | -2.0 (-2.4, -1.7) |
| SGLT2i benefit by 0-3 mmol/mol | 12,935 | -4.2 (-4.3, -4.1) | -2.6 (-2.8, -2.4) |
| GLP1-RA benefit by 0-3 mmol/mol | 13,231 | -4.3 (-4.4, -4.2) | -2.9 (-3.1, -2.7) |
| GLP1-RA benefit by 3-5 mmol/mol | 5,300 | -4.5 (-4.7, -4.3) | -3.3 (-3.6, -3.0) |
| GLP1-RA benefit by ≥5 mmol/mol | 2,914 | -4.4 (-4.7, -4.1) | -3.3 (-3.7, -3.0) |

c) Risk of discontinuation (n=77,741)

| Predicted HbA1c benefit | Risk of discontinuation (%) | | |
|---|---|---|---|
| | N patients | Risk of discontinuation on SGLT2i (95% CI) | Risk of discontinuation on GLP1-RA (95% CI) |
| Overall | 77,741 | 19.8 (19.5, 20.1) | 18.7 (18.1, 19.3) |
| Subgroup | | | |
| SGLT2i benefit by ≥5 mmol/mol | 6,048 | 13.3 (12.0, 14.6) | 16.4 (14.5, 18.4) |
| SGLT2i benefit by 3-5 mmol/mol | 7,657 | 14.3 (13.1, 15.4) | 16.5 (14.7, 18.3) |
| SGLT2i benefit by 0-3 mmol/mol | 23,246 | 18.0 (17.3, 18.7) | 18.6 (17.4, 19.7) |
| GLP1-RA benefit by 0-3 mmol/mol | 24,259 | 20.3 (19.6, 21.0) | 18.7 (17.4, 19.9) |
| GLP1-RA benefit by 3-5 mmol/mol | 10,168 | 22.5 (21.4, 23.7) | 18.0 (16.3, 19.9) |
| GLP1-RA benefit by ≥5 mmol/mol | 6,363 | 26.5 (24.4, 28.7) | 18.9 (16.9, 21.1) |

d) Microvascular complications (n=34,524), with additional adjustment for baseline cardiovascular risk

| Predicted HbA1c benefit | Risk of developing microvascular complications - hazard ratio (less than 1 favours SGLT2i) (95% CI) | | |
|---|---|---|---|
| | N patients | Events | Treatment difference |
| Overall | 34,524 | 5,228 | 0.88 (0.82, 0.94) |
| Subgroup | | | |
| SGLT2i benefit by ≥5 mmol/mol | 3,098 | 561 | 0.81 (0.67, 0.97) |
| SGLT2i benefit by 3-5 mmol/mol | 3,498 | 598 | 0.78 (0.65, 0.94) |
| SGLT2i benefit by 0-3 mmol/mol | 10,288 | 1,655 | 0.87 (0.77, 0.98) |
| GLP1-RA benefit by 0-3 mmol/mol | 11,328 | 1,567 | 0.90 (0.79, 1.03) |
| GLP1-RA benefit by 3-5 mmol/mol | 4,765 | 620 | 0.97 (0.80, 1.18) |
| GLP1-RA benefit by ≥5 mmol/mol | 1,547 | 225 | 1.11 (0.81, 1.53) |

e) Major adverse cardiovascular events (MACE) (n=52,052), with additional adjustment for baseline cardiovascular risk

| Predicted HbA1c benefit | Risk of developing MACE - hazard ratio (less than 1 favours SGLT2i) (95% CI) | | |
|---|---|---|---|
| | N patients | Events | Treatment difference |
| Overall | 52,052 | 1,260 | 1.02 (0.89, 1.18) |
| Subgroup | | | |
| SGLT2i benefit by ≥5 mmol/mol | 4,236 | 80 | 0.69 (0.43, 1.11) |
| SGLT2i benefit by 3-5 mmol/mol | 5,327 | 97 | 0.99 (0.63, 1.57) |
| SGLT2i benefit by 0-3 mmol/mol | 16,279 | 382 | 1.11 (0.86, 1.45) |
| GLP1-RA benefit by 0-3 mmol/mol | 16,419 | 412 | 0.91 (0.71, 1.17) |
| GLP1-RA benefit by 3-5 mmol/mol | 6,763 | 160 | 1.06 (0.70, 1.60) |
| GLP1-RA benefit by ≥5 mmol/mol | 3,028 | 129 | 1.50 (0.92, 2.46) |

f) Heart Failure (n=52,052), with additional adjustment for baseline cardiovascular risk

| Predicted HbA1c benefit | Risk of heart failure - hazard ratio (less than 1 favours SGLT2i) (95% CI) | | |
|---|---|---|---|
| | N patients | Events | Treatment difference |
| Overall | 52,052 | 655 | 0.71 (0.59, 0.85) |
| Subgroup | | | |
| SGLT2i benefit by ≥5 mmol/mol | 4,236 | 42 | 0.96 (0.48, 1.91) |
| SGLT2i benefit by 3-5 mmol/mol | 5,327 | 43 | 0.49 (0.26, 0.90) |
| SGLT2i benefit by 0-3 mmol/mol | 16,279 | 185 | 0.78 (0.55, 1.09) |
| GLP1-RA benefit by 0-3 mmol/mol | 16,419 | 208 | 0.82 (0.58, 1.16) |
| GLP1-RA benefit by 3-5 mmol/mol | 6,763 | 94 | 0.63 (0.39, 1.02) |
| GLP1-RA benefit by ≥5 mmol/mol | 3,028 | 83 | 0.52 (0.33, 0.84) |

g) Developing chronic kidney disease stage 5 or drop of eGFR by 40% (n=52,052), with additional adjustment for baseline cardiovascular risk

| Predicted HbA1c benefit | Risk of chronic kidney disease - hazard ratio (less than 1 favours SGLT2i) (95% CI) | | |
|---|---|---|---|
| | N patients | Events | Treatment difference |
| Overall | 52,052 | 185 | 0.41 (0.30, 0.56) |
| Subgroup | | | |
| SGLT2i benefit by ≥5 mmol/mol | 4,236 | 13 | 0.60 (0.20, 1.83) |
| SGLT2i benefit by 3-5 mmol/mol | 5,327 | 13 | 0.21 (0.07, 0.64) |
| SGLT2i benefit by 0-3 mmol/mol | 16,279 | 42 | 0.30 (0.16, 0.56) |
| GLP1-RA benefit by 0-3 mmol/mol | 16,419 | 53 | 0.65 (0.34, 1.22) |
| GLP1-RA benefit by 3-5 mmol/mol | 6,763 | 36 | 0.31 (0.16, 0.61) |
| GLP1-RA benefit by ≥5 mmol/mol | 3,028 | 28 | 0.50 (0.22, 1.15) |

**sFig. 1**: Variables selected for the prognostic (factors predictive of HbA1c response to SGLT2i therapy) component (A) and the moderator (factors predictive of differential HbA1c response with GLP1-RA compared to SGLT2i therapies) component (B) of the model.

**Posterior inclusion proportions correspond to the average proportion of times each predictor is chosen as a splitting rule divided by the total number of splitting rules appearing in the model component. The threshold used corresponds to one divided by the number of variables used in the model times 100.**

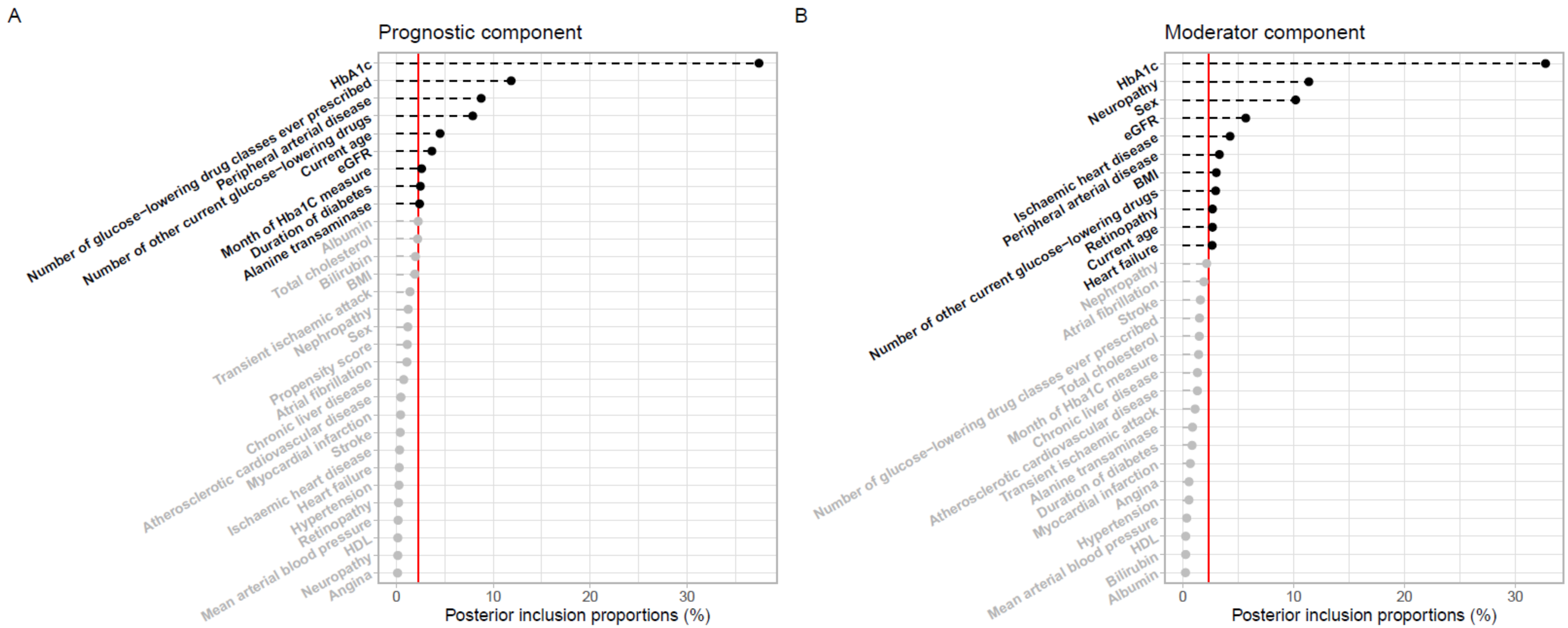

**sFig. 2**: Comparison of predicted outcome HbA1c and predicted conditional average treatment effect (CATE) estimates from two Bayesian Causal Forest (BCF) models with and without including propensity scores in the development cohort.
(A.1) and (A.2) show predictions of outcome HbA1c under both models and a histogram of the predicted HbA1c differences, respectively, and (B.1) and (B.2) are analogous but for the predicted CATE estimates.

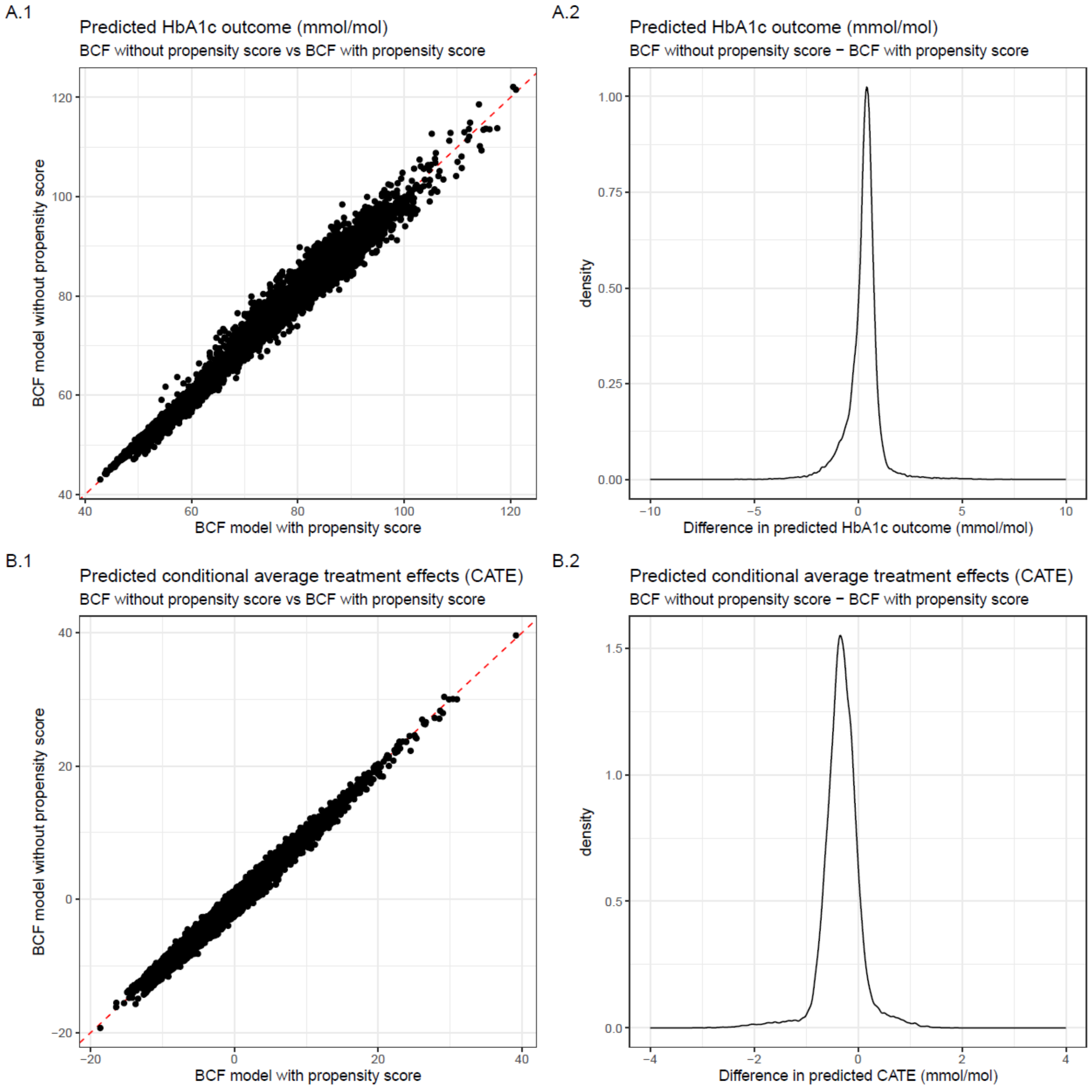

sFig. 3: Calibration plots of predicted conditional treatment effect (CATE) estimates using propensity score matching.

(A) Calibration plots using unadjusted estimates of average treatment effects for each decile of predicted conditional average treatment effects in propensity score matched individuals of the development (A.1) and validation (A.2) cohorts. (B) Calibration plots using estimates adjusted for all variables used in the treatment selection model, in propensity score matched individuals of the development (B.1) and validation (B.2) cohorts.

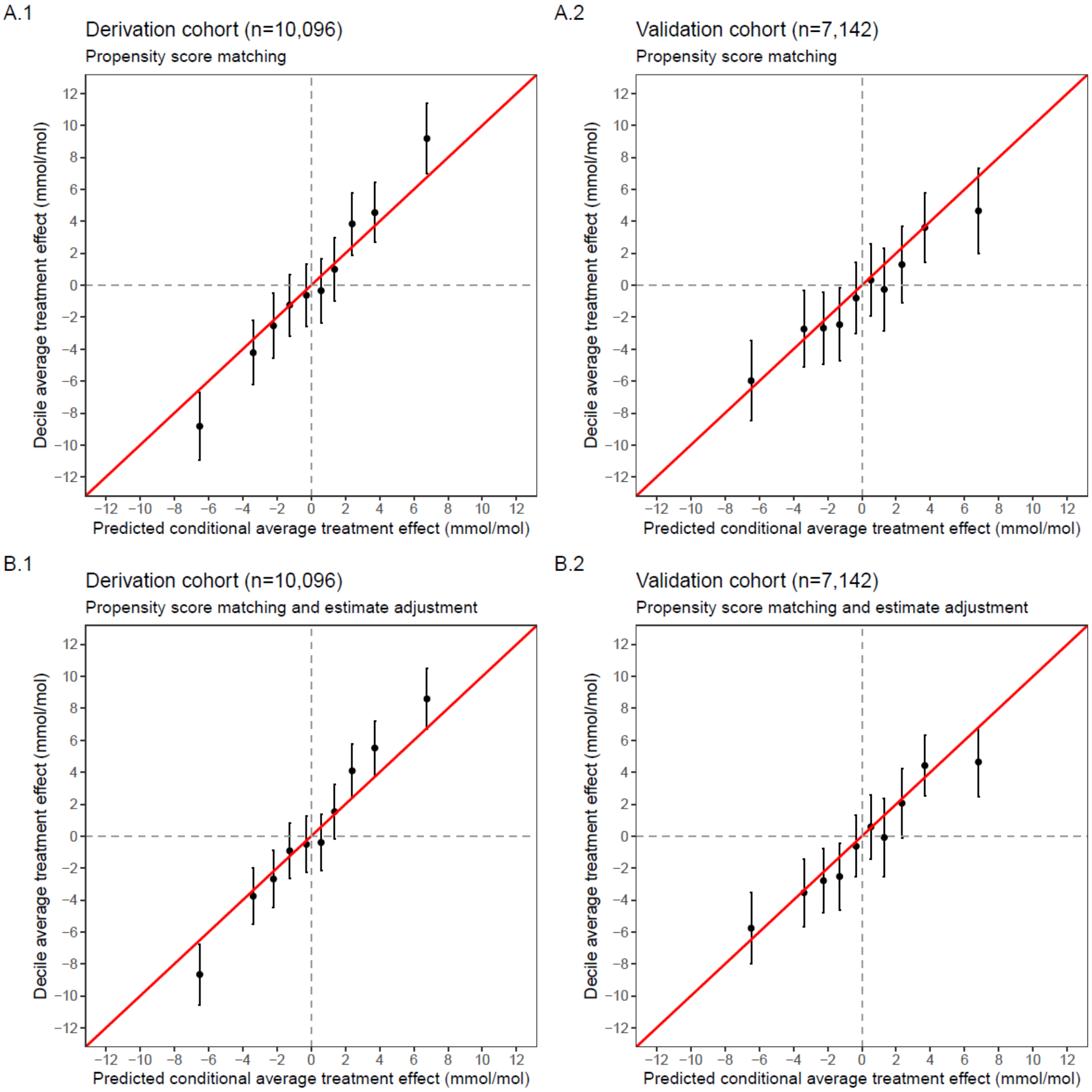

**sFig. 4**: Short-term and long-term clinical outcomes, across subgroups defined by clinical cut-offs of predicted treatment benefit, in propensity score matched cohorts

Replication of Fig. 3 in propensity score matched individuals and without covariate adjustment. Estimates of short-term (A) and long-term (B) outcomes (GLP1-RA baseline group) are calculated with propensity score matched individuals. (A.1) 12-month predicted HbA1c change on each treatment. (A.2) 12-month predicted weight change on each treatment. (A.3) 6-month risk of discontinuation on each treatment. (B.1) 5-year risk of developing microvascular complications. (B.2) 5-year of developing major adverse cardiovascular events (MACE). (B.3) 5-year risk of heart failure.

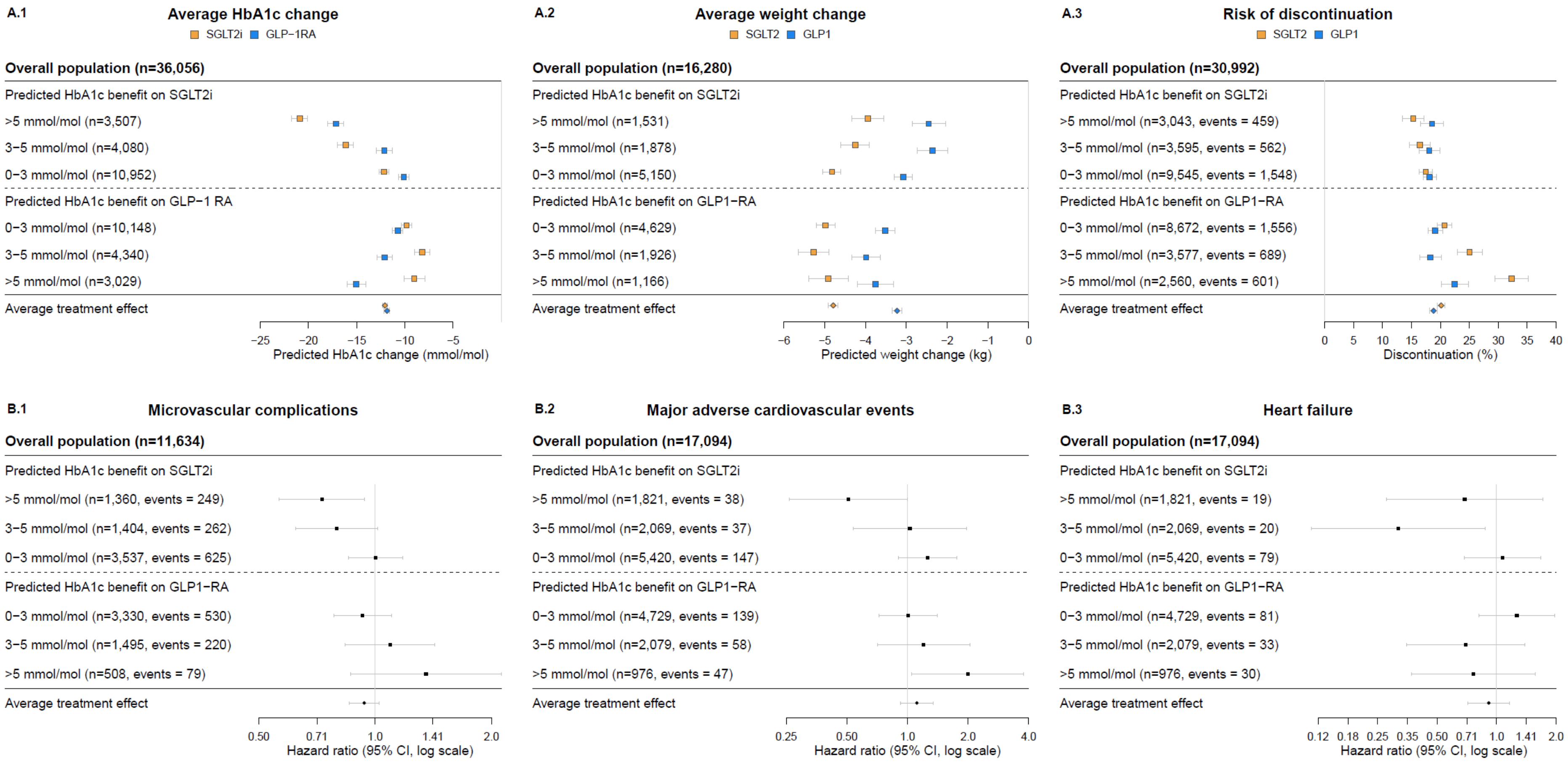

**sFig. 5**: Short-term and long-term clinical outcomes, across subgroups defined by clinical cut-offs of predicted treatment benefit, in propensity score matched cohort with additional covariate adjustment.

Replication of Fig. 3 in propensity score matched individuals. Estimates of short-term (A) and long-term (B) outcomes (GLP1-RA baseline group) are calculated with propensity score matched individuals and adjusted with all the variables used in the treatment selection model. (A.1) 12-month predicted HbA1c change on each treatment. (A.2) 12-month predicted weight change on each treatment. (A.3) 6-month risk of discontinuation on each treatment. (B.1) 5-year risk of developing microvascular complications. (B.2) 5-year of developing major adverse cardiovascular events (MACE). (B.3) 5-year risk of heart failure.

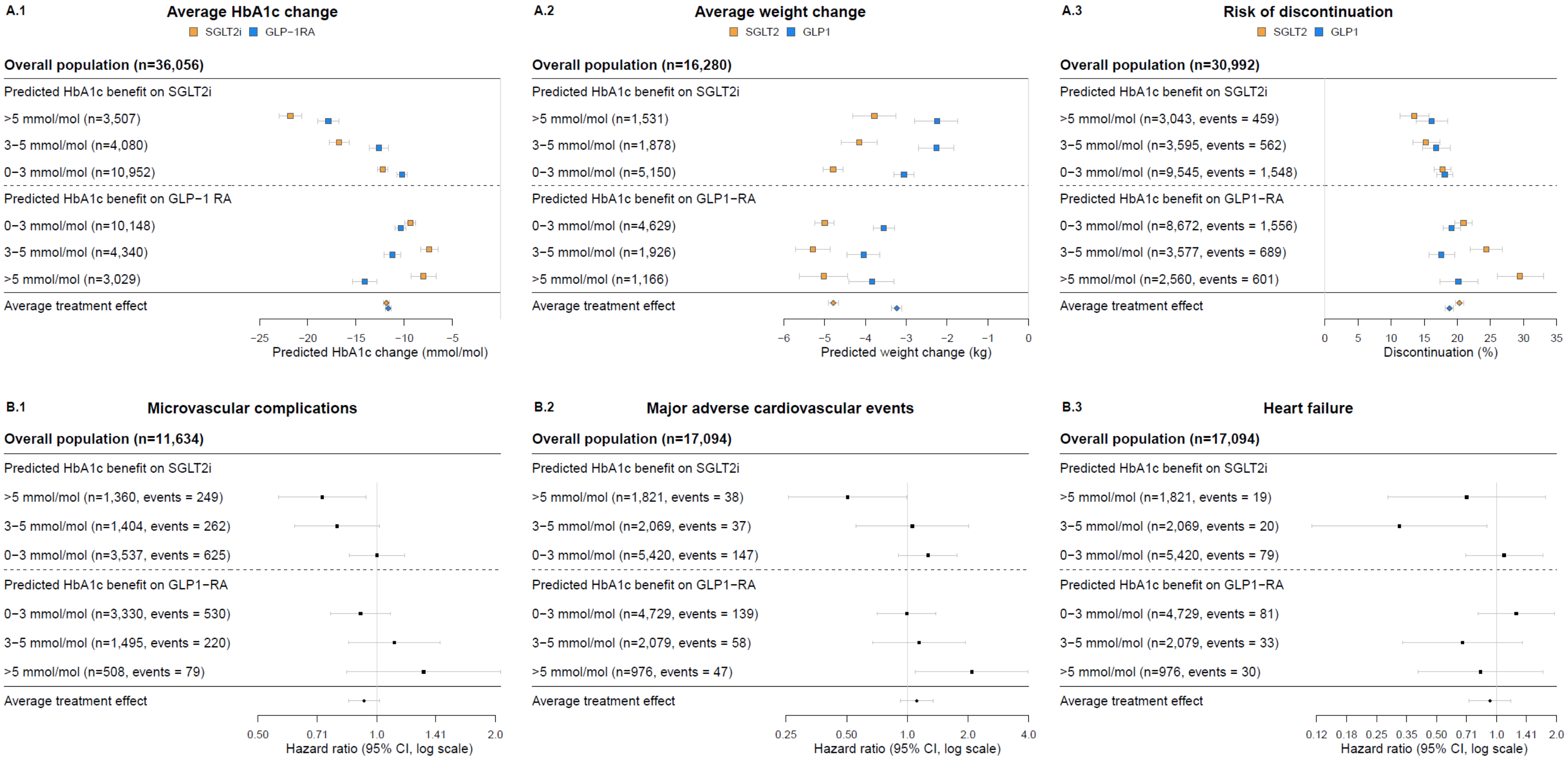

sFig. 6: Relative risk of developing new onset chronic kidney disease (CKD) over 5 years, across subgroups defined by clinical cut-offs of predicted treatment benefit.
Chronic kidney disease (CKD) is defined by a drop of 40% in eGFR or reaching CKD – stage 5. Negative hazard ratio values correspond to a reduced risk of CKD on SGLT2-inhibitor treatment, and positive hazard ratio values correspond to a reduced risk of CKD on GLP-1 receptor agonist treatment. Estimates are split into three approaches: (A) all individuals with estimates adjusted for all the variables used in the treatment selection model; (B) propensity score matched individuals with unadjusted estimates; (C) propensity score matched individuals with estimates adjusted for all the variables used in the treatment selection model.

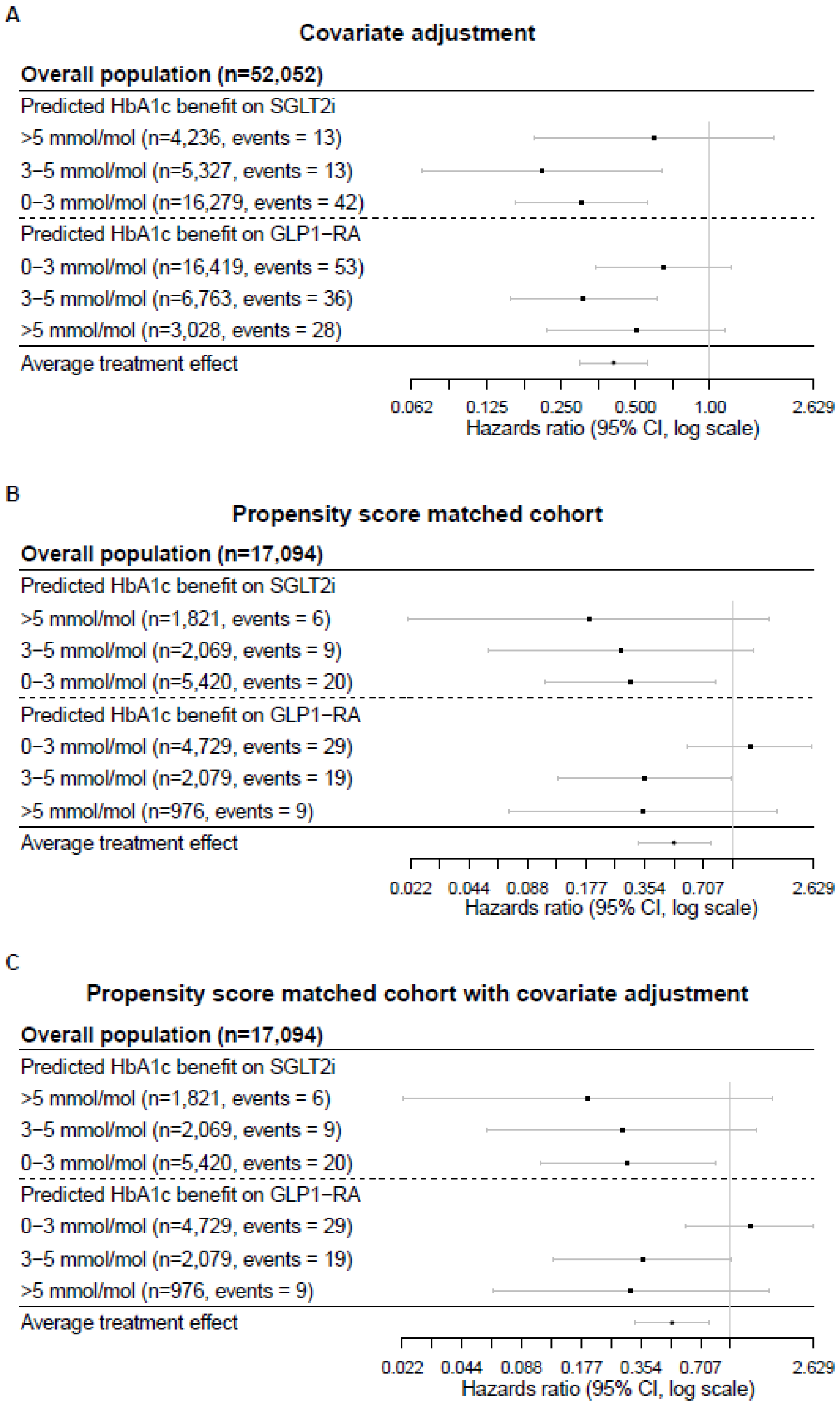

**sFig. 7**: Evaluation of concordance between estimates of predicted HbA1c outcome with SGLT2i from our model and a recently published SGLT2i-DPP4-inhibitor treatment selection model [1].

82,933 eligible individuals from the overall study population fulfilling inclusion criteria for both studies. (A) Comparison of outcome HbA1c predictions for all patients from both models. The red dashed line corresponds to a theoretical equality between both models. The blue line corresponds to the trend line of the predictions. (B) Histogram of residual difference between HbA1c predictions (mmol/mol) of both models.

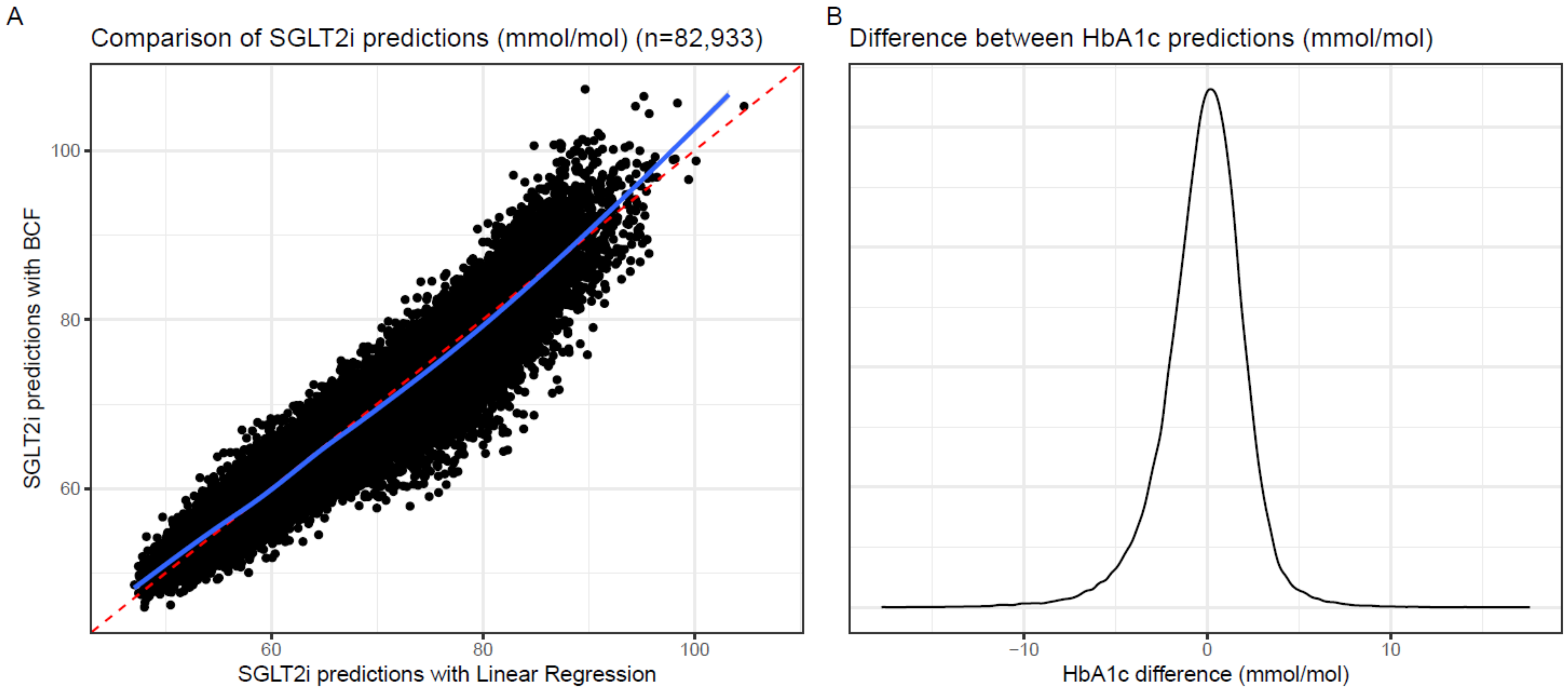

sFig. 8: Variable selection plot for the propensity score model — identifying the most important predictors.
This plot presents the variable selection results of a propensity score model. The green lines represent the threshold levels determined from permutation distributions that must be exceeded for a variable to be selected by the model. The plotted points indicate the variable inclusion proportions for the observed data in the propensity score model. The solid dots represent the variables that are included in the model because their observed values exceed the corresponding green bars, while the open dots represent the variables that are not included in the model because their observed values fall below the threshold.

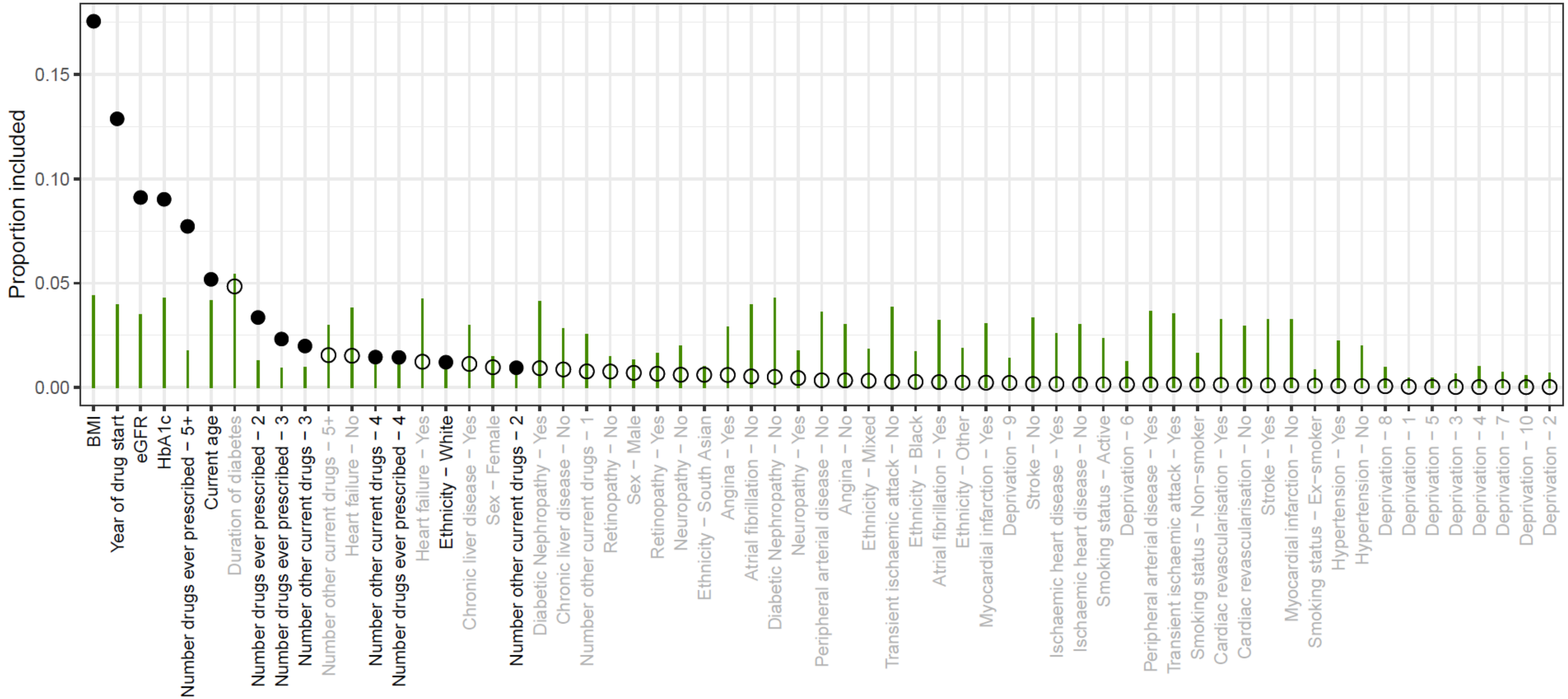

sFig. 9: Received operating characteristic (ROC) and precision-recall curves of the propensity score model developed in the derivation cohort and validated in hold-out validation data.

The ROC curves (1) see similar performance in the derivation (A) and validation (B) cohorts, showing the model is performing well in correctly identifying when SGLT2i and GLP-1RA are indicated. Precision-recall curves (2) plot the proportion of true positives among all predicted positives, against the proportion of true positives among all actual positives. The precision-recall curves see similar performance in the derivation and validation cohorts, showing that the model can accurately identify positive instances (GLP-1RA) with minimal false positives for a proportion of thresholds.

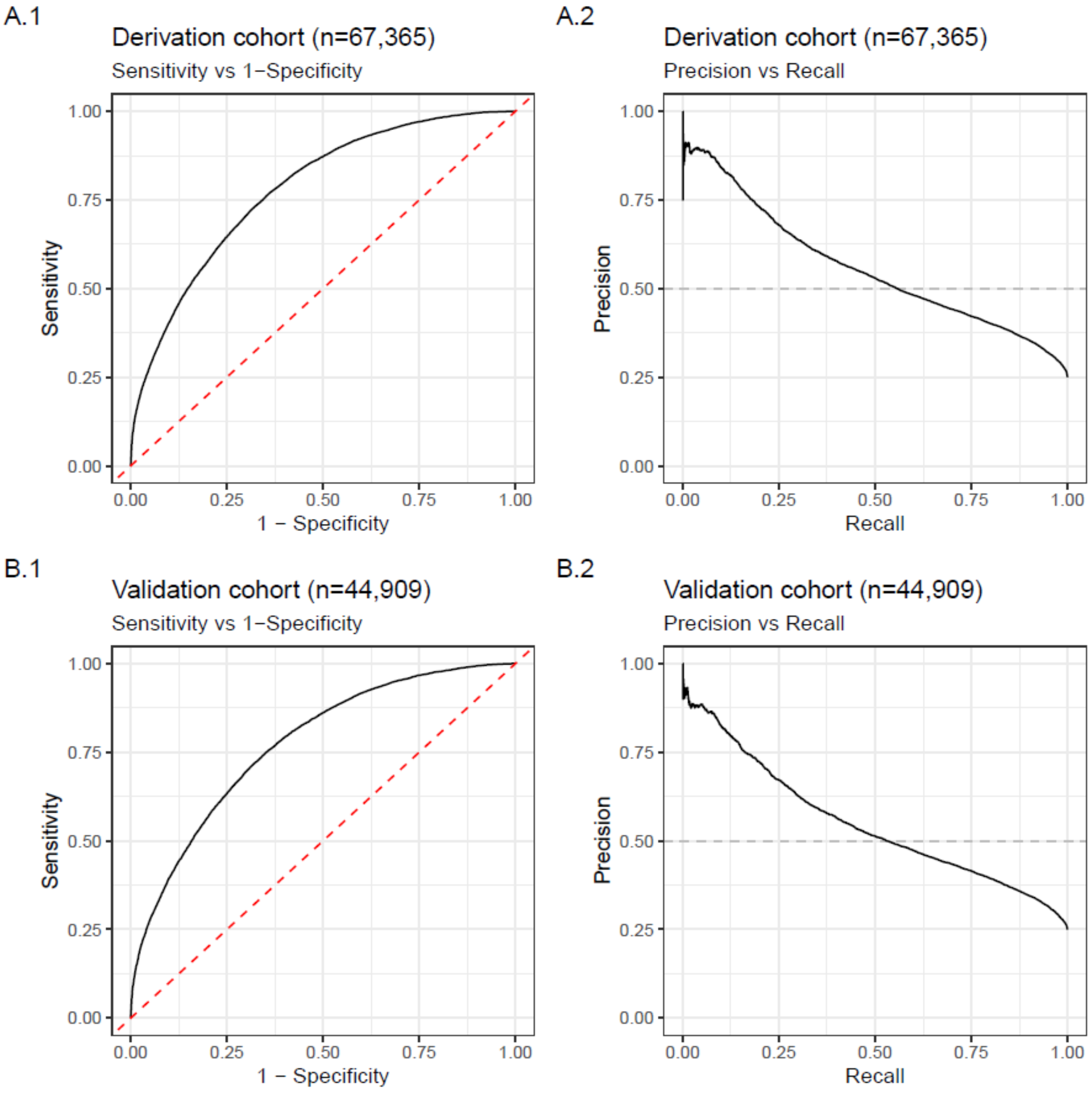

sFig. 10: Calibration plots of predicted conditional individualised treatment effects in the validation cohort for those with and without cardiovascular disease (CVD).
Calibration in those with (A) and without (B) CVD in the hold-out validation cohort. Estimates are adjusted with all the variables used in the treatment selection model.

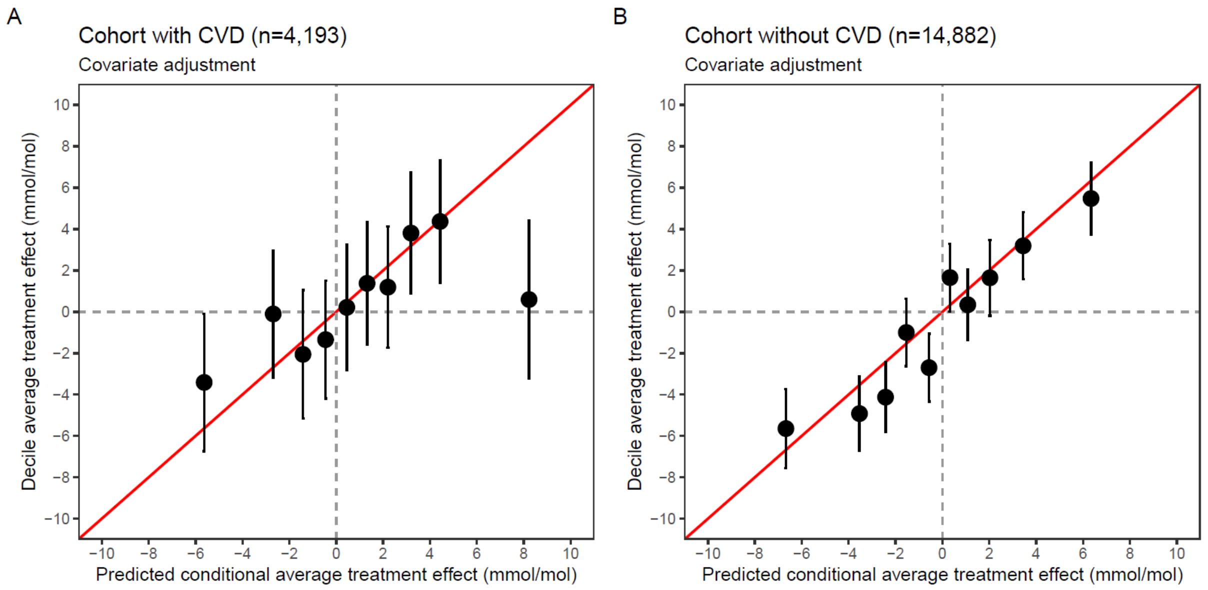

sFig. 11: Comparison of predicted outcome HbA1c and predicted conditional average treatment effect (CATE) estimates from a sparse Bayesian Causal Forest (BCF) model and a BCF model with different sets of variables. Sparse BCF is fitted with all candidate predictors, and BCF is fitted with the selected variables from variable selection (see Table 2). (A.1) and (A.2) show predictions of outcomes HbA1c under both models and a histogram of the predicted HbA1c differences, respectively, and (B.1) and (B.2) are analogous but for the predicted CATE estimates.

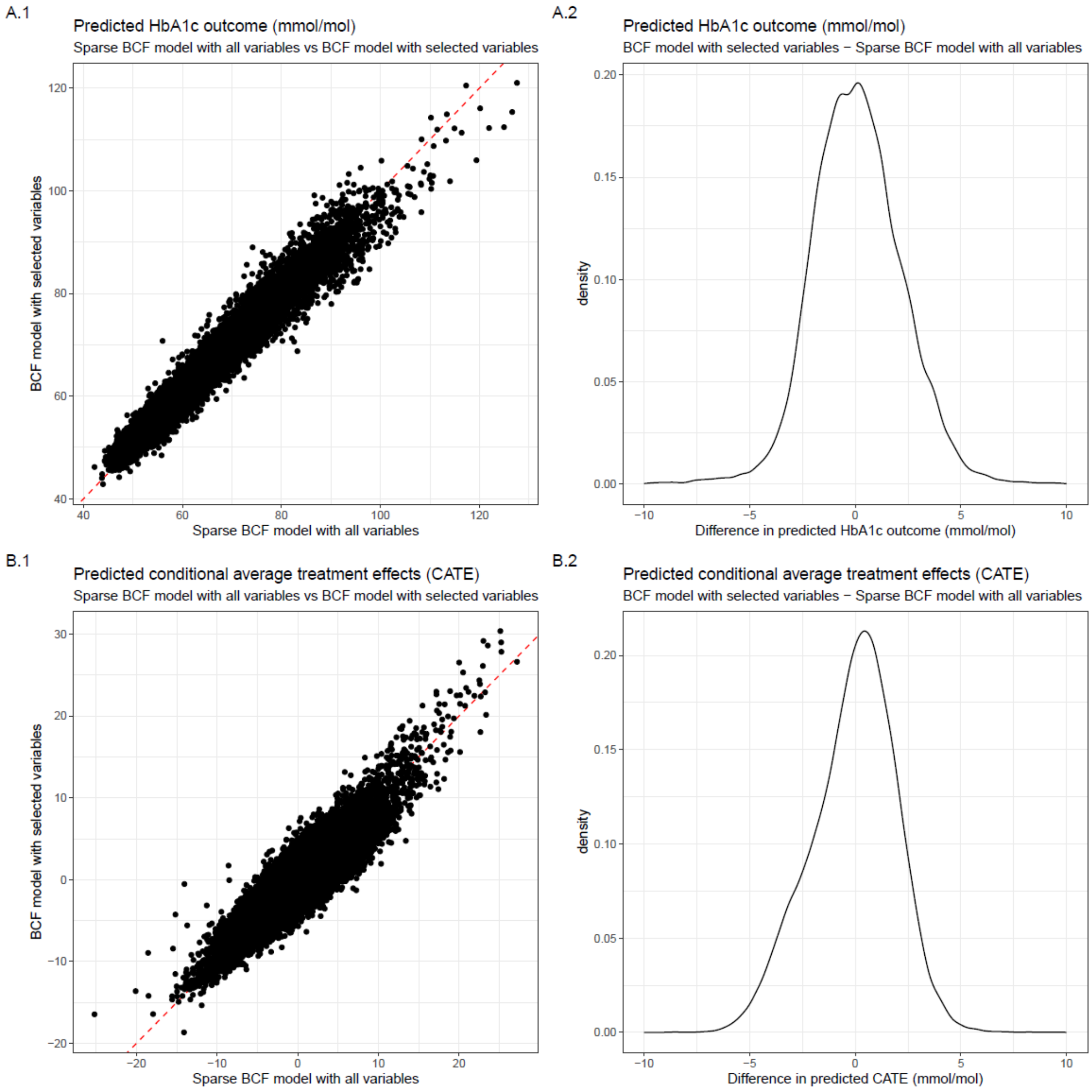

| Section/Topic | Item | | Checklist Item | Page |
|---|---|---|---|---|
| **Title and abstract** | | | | |
| Title | 1 | D;V | Identify the study as developing and/or validating a multivariable prediction model, the target population, and the outcome to be predicted. | 1 |
| Abstract | 2 | D;V | Provide a summary of objectives, study design, setting, participants, sample size, predictors, outcome, statistical analysis, results, and conclusions. | 1 |
| **Introduction** | | | | |
| Background and objectives | 3a | D;V | Explain the medical context (including whether diagnostic or prognostic) and rationale for developing or validating the multivariable prediction model, including references to existing models. | 2 |
| | 3b | D;V | Specify the objectives, including whether the study describes the development or validation of the model or both. | 2 |
| **Methods** | | | | |
| Source of data | 4a | D;V | Describe the study design or source of data (e.g., randomized trial, cohort, or registry data), separately for the development and validation data sets, if applicable. | 3 |
| | 4b | D;V | Specify the key study dates, including start of accrual; end of accrual; and, if applicable, end of follow-up. | 3 |
| Participants | 5a | D;V | Specify key elements of the study setting (e.g., primary care, secondary care, general population) including number and location of centres. | 3 |
| | 5b | D;V | Describe eligibility criteria for participants. | 3,8 |
| | 5c | D;V | Give details of treatments received, if relevant. | 3 |
| Outcome | 6a | D;V | Clearly define the outcome that is predicted by the prediction model, including how and when assessed. | 3,8 |
| | 6b | D;V | Report any actions to blind assessment of the outcome to be predicted. | 8 |
| Predictors | 7a | D;V | Clearly define all predictors used in developing or validating the multivariable prediction model, including how and when they were measured. | 8,9 |
| | 7b | D;V | Report any actions to blind assessment of predictors for the outcome and other predictors. | 8 |
| Sample size | 8 | D;V | Explain how the study size was arrived at. | 3,8 |
| Missing data | 9 | D;V | Describe how missing data were handled (e.g., complete-case analysis, single imputation, multiple imputation) with details of any imputation method. | 3 |
| Statistical analysis methods | 10a | D | Describe how predictors were handled in the analyses. | 3,8 |
| | 10b | D | Specify type of model, all model-building procedures (including any predictor selection), and method for internal validation. | 3,8,9,10 |
| | 10c | V | For validation, describe how the predictions were calculated. | 10,11 |
| | 10d | D;V | Specify all measures used to assess model performance and, if relevant, to compare multiple models. | 10,11 |
| | 10e | V | Describe any model updating (e.g., recalibration) arising from the validation, if done. | - |
| Risk groups | 11 | D;V | Provide details on how risk groups were created, if done. | - |
| Development vs. validation | 12 | V | For validation, identify any differences from the development data in setting, eligibility criteria, outcome, and predictors. | 3 |
| **Results** | | | | |
| Participants | 13a | D;V | Describe the flow of participants through the study, including the number of participants with and without the outcome and, if applicable, a summary of the follow-up time. A diagram may be helpful. | 3 |
| | 13b | D;V | Describe the characteristics of the participants (basic demographics, clinical features, available predictors), including the number of participants with missing data for predictors and outcome. | 3 |
| | 13c | V | For validation, show a comparison with the development data of the distribution of important variables (demographics, predictors and outcome). | 3 |
| Model development | 14a | D | Specify the number of participants and outcome events in each analysis. | 3,4 |
| | 14b | D | If done, report the unadjusted association between each candidate predictor and outcome. | - |
| Model specification | 15a | D | Present the full prediction model to allow predictions for individuals (i.e., all regression coefficients, and model intercept or baseline survival at a given time point). | - |
| | 15b | D | Explain how to the use the prediction model. | 8,9,10 |
| Model performance | 16 | D;V | Report performance measures (with CIs) for the prediction model. | 3,4 |
| Model-updating | 17 | V | If done, report the results from any model updating (i.e., model specification, model performance). | 3,4 |
| **Discussion** | | | | |
| Limitations | 18 | D;V | Discuss any limitations of the study (such as nonrepresentative sample, few events per predictor, missing data). | 6,7 |
| Interpretation | 19a | V | For validation, discuss the results with reference to performance in the development data, and any other validation data. | 6,7 |
| | 19b | D;V | Give an overall interpretation of the results, considering objectives, limitations, results from similar studies, and other relevant evidence. | 6,7 |
| Implications | 20 | D;V | Discuss the potential clinical use of the model and implications for future research. | 6,7 |
| **Other information** | | | | |
| Supplementary information | 21 | D;V | Provide information about the availability of supplementary resources, such as study protocol, Web calculator, and data sets. | 5,9 |
| Funding | 22 | D;V | Give the source of funding and the role of the funders for the present study. | 12 |

*Items relevant only to the development of a prediction model are denoted by D, items relating solely to a validation of a prediction model are denoted by V, and items relating to both are denoted D;V. We recommend using the TRIPOD Checklist in conjunction with the TRIPOD Explanation and Elaboration document.

### Priors of the model.

#### Propensity score model BART

The Bayesian additive regression trees (BART) technique employs a Bayesian methodology for non-parametric function estimation through regression trees [2]. These trees employ a recursive binary partitioning of the predictor space, creating a series of hyperrectangles to approximate an unknown function $f$. The predictor space's dimension equals the number of variables, $p$. Due to their flexible fitting of interactions and non-linearities, regression tree models are helpful. Models composed of multiple regression trees can capture interactions, non-linearities, and additive effects in $f$.

BART can be considered a sum-of-trees ensemble, with a novel estimation approach relying on a fully Bayesian probability model. Specifically, the BART model can be expressed as:

$$\boldsymbol{Y} = f(\boldsymbol{X}) + \boldsymbol{\varepsilon} \approx \boldsymbol{T}_1^{\boldsymbol{M}}(\boldsymbol{X}) + \boldsymbol{T}_2^{\boldsymbol{M}}(\boldsymbol{X}) + \cdots + T_m^M(\boldsymbol{X}) + \boldsymbol{\varepsilon}, \qquad \boldsymbol{\varepsilon} \sim \boldsymbol{N}_n\left(\boldsymbol{0}, \boldsymbol{\sigma}^2 \boldsymbol{I}_n\right) \tag{1}$$

In this context, $\boldsymbol{Y}$ represents the response vector with dimensions $n \times 1$, while $\boldsymbol{X}$ is the $n \times p$ design matrix with column-joined predictors. Additionally, $\boldsymbol{\varepsilon}$ represents the $n \times 1$ noise vector. The analysis involves $m$ separate regression trees, each consisting of a tree structure (referred to as $\mathbf{T}$) and the parameters at the terminal nodes or leaves (referred to as $\mathbf{M}$). When combined, both components make up the complete tree denoted as $\mathbf{T}^{\mathbf{M}}$. This complete representation encompasses both the structure and set of leaf parameters.

In the BART model, the prior consists of three distinct components. Firstly, it includes the tree structure itself. Secondly, it incorporates the leaf parameters given the tree structure. Finally, it encompasses the error variance $\boldsymbol{\sigma}^2$, independent of the tree structure and leaf parameter.

$$\mathbb{P}(\mathbf{T}_1^{\mathbf{M}}, \dots, \mathbf{T}_m^{\mathbf{M}}, \boldsymbol{\sigma}^2) = \left[\prod_t \mathbb{P}(\mathbf{T}_t^{\mathbf{M}})\right] \mathbb{P}(\boldsymbol{\sigma}^2) = \left[\prod_t \mathbb{P}(\mathbf{M}_t | \mathbf{T}_t) \mathbb{P}(\mathbf{T}_t)\right] \mathbb{P}(\boldsymbol{\sigma}^2)$$
$$= \left[\prod_t \prod_l \mathbb{P}(\boldsymbol{\mu}_{t,l} | \mathbf{T}_t) \mathbb{P}(\mathbf{T}_t)\, \mathbb{P}(\boldsymbol{\sigma}^2)\right]$$

Initially, we discuss $\mathbb{P}(\mathbf{T}_t)$, a component of the prior that impacts node placement within the tree. The probability of nonterminal nodes at a particular depth $d$ is $\alpha(1+d)^{-\beta}$ where $\alpha \in (0,1)$ and $\beta \in [0, \infty]$. In this context, depth is measured as the distance from the root. For instance, the root node has a depth of 0, while its first child node has a depth of 1, and so on. This prior form can limit the complexity of any single tree by enforcing shallow tree structures. The values for these hyperparameters are $\alpha = 0.95$ and $\beta = 2$, in line with Chipman *et al.* [2].

Next, we discuss the prior component $\mathbb{P}(\mathbf{M}_t | \mathbf{T}_t)$ which regulates the leaf parameters. When a tree has a collection of terminal nodes, each node (or leaf) has a continuous parameter representing the "best guess" of the response in that specific partition of predictor space. This parameter is the fitted value assigned to any observation within that node. The prior on each of the leaf parameters is $\boldsymbol{\mu}_l \sim^{iid} \boldsymbol{N}(\boldsymbol{\mu}_\mu / m, \boldsymbol{\sigma}_\mu^2)$, where the expectation, $\boldsymbol{\mu}_\mu$, is selected as the range centre, specifically $(y_{min} + y_{max})/2$. However, outliers can impact the range centre, which can be remedied by logging the response or applying windsorisation.

The variance is chosen empirically such that the range centre plus or minus $k = 2$ variances cover 95% of the response values provided in the training set (where $k = 2$ corresponds to 95% coverage). Therefore, given $m$ trees, we automatically employ $\boldsymbol{\sigma}_\mu$ such that $m\boldsymbol{\mu}_\mu - k\sqrt{m}\boldsymbol{\sigma}_\mu = y_{min}$ and $m\boldsymbol{\mu}_\mu +$

$k\sqrt{m}\boldsymbol{\sigma}_{\boldsymbol{\mu}} = y_{max}$. This prior aims to achieve model regularisation by shrinking the leaf parameters towards the centre of the response distribution.

The last prior is on the error variance, and it is selected as $\boldsymbol{\sigma}^2 \sim InvGamma(\nu/2, \nu\lambda/2)$. $\lambda$ is calculated from the data to ensure that there is a $q = 90\%$ probability that the BART model will outperform the RMSE of an ordinary least squares regression. Consequently, the majority of the prior probability mass is positioned beneath the RMSE from the least squares regression. Furthermore, this prior restricts the likelihood of small $\boldsymbol{\sigma}^2$ values to prevent overfitting.

To generate draws from the posterior distribution of $\mathbb{P}\left(\mathbf{T}_1^{\mathbf{M}}, \dots, \mathbf{T}_m^{\mathbf{M}}, \boldsymbol{\sigma}^2 \middle| \boldsymbol{\mu}\right)$, a Gibbs sampler is utilised. A key feature of the Gibbs sampler for BART is to employ a form of "Bayesian backfitting" where the $j^{\text{th}}$ tree is fit iteratively, holding all other $m - 1$ trees constant by exposing only the residual response that remains unfitted. This relies on Metropolis-Hastings draws from the posterior of the tree distributions, which involve introducing minor perturbations to the tree structure, such as growing a terminal node by adding two child nodes, pruning two child notes to render their parent node terminal, or modifying a split rule. These three potential tree adjustments are labelled as GROW, PRUNE, AND CHANGE. The prior probabilities for proposing changes to the tree structures are: GROW $= 2.5/9$, PRUNE $= 2.5/9$ and CHANGE $= 4/9$.

BART was modified to handle classification problems for categorical response variables. For the binary classification problem (coded with outcomes "0" and "1"), we assume a probit model:

$$\mathbb{P}(\boldsymbol{Y} = 1|\boldsymbol{X}) = \boldsymbol{\Phi}\left(\mathbf{T}_1^{\mathbf{M}}(\boldsymbol{X}) + \mathbf{T}_2^{\mathbf{M}}(\boldsymbol{X}) + \dots + \mathbf{T}_m^{\mathbf{M}}(\boldsymbol{X})\right),$$

where $\boldsymbol{\Phi}$ denotes the cumulative density function of the standard normal distribution. In this formulation, the sum-of-trees model serves as an estimate of the conditional probit at $\boldsymbol{x}$**,** which can be easily transformed into a conditional probability estimate of $Y = 1$. The prior on $\boldsymbol{\sigma}^{\mathbf{2}}$ is not needed in the classification setting as the model assumes $\boldsymbol{\sigma}^2 = 1$. The prior on the tree structure remains the same as in the regressions setting, and a few minor modifications are required for the prior on the leaf parameters. See Chipman *et al.* (2010) [2] and Kapelner *et al.* (2013) [3] for more details.

#### Sparse BCF

Bayesian Additive Regression Trees (BART) are a non-parametric regression model used to estimate conditional expectations of a response variables $Y_i$ via a "sum-of-trees". We can redefine equation (1) to be:

$$Y_i = f(\boldsymbol{X}_i, Z_i) + \varepsilon_i, \quad where\ \varepsilon_i \sim N(0, \sigma^2)$$

Considering this regression framework, we can use BART to flexibly represent $f(\,.\,)$ as

$$f(\boldsymbol{X}, Z) = \sum_{j=1}^{m} g_j\left([\boldsymbol{X}\ \ Z], (T_j, M_j)\right),$$

where $m$ is the total number of trees in the model, and each tree is defined by a pair $(T_j, M_j)$. $T_j$ represents the binary split rules of the tree, while $M_j$ is the set of terminal nodes in that tree. The function $g_j(\,.\,)$ is specific to each tree and maps the predictors $[\boldsymbol{X}\ \ Z]$ to the set of terminal nodes $M_j$, following the binary split rules in $T_j$. The conditional mean function $f(\boldsymbol{x}, z) = \mathbb{E}[Y_i|\boldsymbol{X}_i = \boldsymbol{x}_i, Z_i = z_i]$ is calculated by adding up all the terminal nodes $\psi_{ij}$ assigned to the predictors $[\boldsymbol{X}\ \ Z]$ by the tree functions $g_j(\,.\,)$, that is $\sum_{j=1}^{m} g_j(\,.\,)$ [4].

This BART method uses a two-stage regression approach:

$$Z_i \sim Bernoulli\left(\pi(\widetilde{\boldsymbol{X}}_i)\right), \pi(\widetilde{\boldsymbol{x}}_i) = \mathbb{P}(Z_i = 1|\widetilde{\boldsymbol{X}}_i = \widetilde{\boldsymbol{x}}_i) \tag{2}$$

$$Y_i = \mu([\boldsymbol{X}_i \ \pi(\widetilde{\boldsymbol{X}}_i)]) + \tau(\boldsymbol{W}_i)Z_i + \varepsilon_i \tag{3}$$

The equation (2) deals with propensity score estimation. Equation (3) estimates the prognostic score $\mu(\,.\,)$, defined as the effect of the covariates $\boldsymbol{X}_i \in \mathrm{X}$ on the outcome $Y_i$ in the absence of treatment $\mu(\boldsymbol{x}_i) = \mathbb{E}[Y_i|\boldsymbol{X}_i = \boldsymbol{x}_i, Z_i = 0]$, and CATE $\tau(\,.\,)$.

BART and BCF can effectively handle sparsity due to the random uniform selection of splitting variables. However, they do not explicitly incorporate heterogeneous sparsity or feature shrinkage, which results in the assumption of equal levels of heterogeneity for all covariates included in the model. Let us define first $\boldsymbol{s} = (s_1, \dots, s_p)$ as the vector of splitting probabilities of each predictor $j \in \{1, \dots, P\}$, where each $s_j$ represents the probability for the $j$th predictor of being chosen as a splitting variable in one of the decision nodes of a tree. Shrinkage BCF places symmetric Dirichlet priors in the estimation of prognostic $\mu(\,.\,)$ and moderating effects $\tau(\,.\,)$ to induce sparsity. For now, we will only consider the case where $\boldsymbol{W}_i = \boldsymbol{X}_i$, that is, where the same set of covariates is used for the estimation of $\mu(\,.\,)$ and $\tau(\,.\,)$. The priors are:

$$\boldsymbol{s}_\mu \sim \text{Dirichlet}\left(\frac{\alpha_\mu}{P+1}, \dots, \frac{\alpha_\mu}{P+1}\right), \qquad \frac{\alpha_\mu}{\alpha_\mu + \rho_{mu}} \sim \text{Beta}(a, b)$$

$$\boldsymbol{s}_\tau \sim \text{Dirichlet}\left(\frac{\alpha_\tau}{P}, \dots, \frac{\alpha_\tau}{P}\right), \qquad \frac{\alpha_\tau}{\alpha_\tau + \rho_\tau} \sim \text{Beta}(a, b)$$

where the Beta's parameters are chosen to be $(a, b) = (0.5, 1)$. The hyperparameter $\rho$ is set equal to $(P + 1)$ in the case of the prognostic score, and it is set to $\frac{P}{2}$ in the case of the moderator effect.

The values for leaf hyperparameters are $\alpha = 0.95$ and $\beta = 2$. The tree structure hyperparameters for the prognostic and moderator effects are similar, corresponding to a sum of 200 trees, leaf hyperparameters $= 0.95$ and $\beta = 2$, $\boldsymbol{\sigma}^2 = 2 \times SD(Y_i)$ (standard deviation [SD]). See Caron *et al.* (2021) for more details.

#### BCF – treatment selection model

For the treatment selection model, we revert the prior on splitting probabilities to a uniform distribution. Otherwise, everything is similar to everything mentioned in the previous sections [5].

#### Additional outcomes

HbA1c and weight outcomes are modelled with Bayesian linear regressions, placing a normal distribution of mean 0 and variance 2 on all parameters. Discontinuation is modelled with a Bayesian logistic regression model, placing a normal distribution of mean 0 and a variance 2 on all parameters.

Long-term outcomes are modelled with a Bayesian survival model, placing a normal distribution of mean 0 and variance 2 on all parameters.
